# Single Cell Characterization of Myeloma and its Precursor Conditions Reveals Transcriptional Signatures of Early Tumorigenesis

**DOI:** 10.1101/2022.02.01.22270128

**Authors:** Rebecca Boiarsky, Nicholas J. Haradhvala, Jean-Baptiste Alberge, Romanos Sklavenitis-Pistofidis, Tarek H Mouhieddine, Oksana Zavidij, Ming-Chieh Shih, Danielle Firer, Mendy Miller, Habib El-Khoury, Shankara K. Anand, François Aguet, David Sontag, Irene M. Ghobrial, Gad Getz

**Author notes:** ***Corresponding Author(s):*** David Sontag, PhD, Massachusetts Institute of Technology, 77 Massachusetts Ave, Cambridge, MA 02139, Irene M. Ghobrial, MD, Medical Oncology, Dana-Farber Cancer Institute, 450 Brookline Ave, Boston, MA 02215, Gad Getz, PhD, Broad Institute of MIT and Harvard, 75 Ames St., Cambridge, MA 02142. These authors jointly supervised this work (David Sontag, Irene M. Ghobrial, Gad Getz).

## Abstract

Multiple myeloma is a plasma cell malignancy almost always preceded by precursor conditions, but low tumor burden of these early stages has hindered the study of their molecular programs through bulk sequencing technologies. Here, we generated and analyzed single cell RNA-sequencing of plasma cells from 26 patients at varying disease stages and 9 healthy donors. In silico dissection and comparison of normal and transformed plasma cells from the same bone marrow biopsy enabled discovery of novel, patient-specific transcriptional changes. Using Bayesian Non-Negative Matrix Factorization, we discovered 15 gene expression signatures which represent transcriptional modules relevant to myeloma biology, and identified a signature that is uniformly lost in neoplastic cells across disease stages. Finally, we demonstrated that tumors contain heterogeneous subpopulations expressing distinct transcriptional patterns. Our findings characterize transcriptomic alterations present at the earliest stages of myeloma, paving the way for exploration of personalized treatment approaches prior to malignant disease.

## Main

Multiple myeloma (MM) is a plasma cell (PC) malignancy residing in the bone marrow (BM)^1^. MM is almost always preceded by the precursor states monoclonal gammopathy of undetermined significance (MGUS) and smoldering multiple myeloma (SMM)^1–3^. However, the progression risk is highly heterogeneous, whereby certain patients progress quickly, while others never do. Patients with SMM exhibit progression rates of 10% per year, compared to just 1% for MGUS^4, 5^. Currently, our ability to predict progression is based only on a few clinical parameters (e.g., M-spike, light chains, and percent tumor burden)^6–8^. Therefore, there is a need to further define molecular characteristics of patients who are at risk of progression. A thorough characterization of pre-malignant cells and the state of the microenvironment in MGUS and SMM patients can help us distinguish the molecular mechanisms that underlie initial tumorigenesis versus later progression, predict which individuals are most at risk for progression, and identify potential targets for early therapeutic intervention.

Our understanding of genetic changes associated with disease progression and tumor evolution in MM is founded on studies that use bulk analysis, including microarrays and DNA sequencing. It has been shown that MGUS and SMM clones may already harbor chromosomal alterations that define MM (translocations involving IgH or hyperdiploidy)^2, 7^, and progression in MM is driven by the acquisition of events like *MYC* translocations, chromosome 1q gains/amplifications and *TP53* mutations^9, 10^.

At the RNA level, it is challenging to draw conclusions about the phenotype of pre-malignant cells and the dynamics of malignant transformation from bulk RNA-sequencing studies^11^ due to low tumor purity (i.e., fraction of tumor cells in a sample) at the precursor stages. Recently, single cell studies of precursor conditions^12, 13^ have allowed for characterization of pre-malignant cells, but such datasets are still scarce and require careful computational analysis in order to glean insights from the limited number of neoplastic cells present in biopsies from patients with precursor disease.

In this study, we generated and analyzed single cell RNA-sequencing (scRNA-seq) data from 29,387 PCs representing 26 samples from patients with MGUS, SMM, or MM as well as 9 normal bone marrow donors (NBM). The single cell resolution of our data and the fact that those precursor samples contain a mixture of neoplastic and normal cells allowed us to isolate the transcriptional changes of neoplastic cells compared to normal plasma cells within the same patient sample and to characterize their transcriptomes across the disease spectrum. We have previously analyzed the immune microenvironment of these same patients^14^, and here we explore transcriptional changes within tumor cells as well as correlations between tumor and immune cell activity in our cohort. We employed novel methods for identifying neoplastic cells from within a mixed sample, report our findings from a nuanced within-patient differential expression analysis approach, and employ Bayesian non-negative matrix factorization (NMF) to highlight gene signatures that are active in our cohort and validated in external cohorts. Taken together, our study (i) presents a highly detailed and comprehensive view of the transcriptional transformation occurring in individual patients with myeloma and its precursor conditions, (ii) discovers gene expression signatures that are shared across patients with different driver events and at different stages of disease, and (iii) characterizes heterogeneity both between and within tumors.

## Results

### Single cell transcriptional profiles reflect driver events and reveal patient-specific patterns

To investigate the gene expression dynamics of PCs at different stages of MM progression, we performed droplet-based single-cell RNA sequencing of 35 samples isolated from BM aspirates of patients with MGUS (n=6), SMM (n=12), and newly diagnosed MM (n=8), as well as nine healthy donors (NBM, n=9; Fig. 1a; Supplementary Tables 1 and 2). One patient was biopsied both at the SMM stage and after progression to MM (SMM-1 and MM-8).

**Figure 1.**
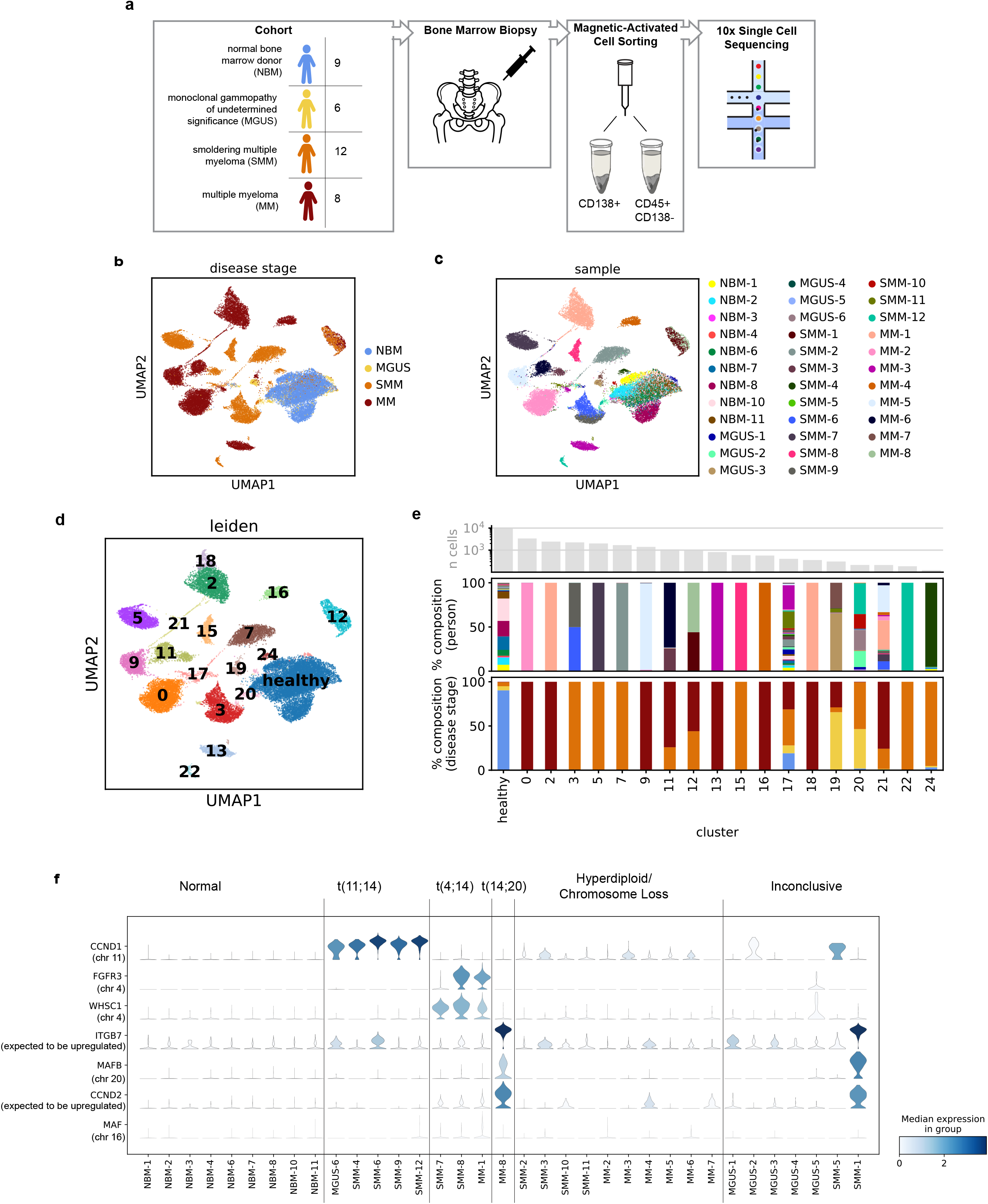
The landscape of healthy and malignant plasma cells at single cell resolution. **a,** Overview of cohort and experimental setup. **a,b,** UMAP representation of plasma cells colored by disease stage **(b)** and sample ID **(c)**. Cells similar in expression profile are placed nearby in this embedding. **d,** Results of leiden clustering of all cells. Seven clusters were merged to define a single cluster of normal plasma cells. **e,** Sample composition of leiden clusters, by disease stage and sample ID (colors match the legends given in (**b)** and (**c**), respectively). The majority of clusters each consist of cells from a single sample. **f,** Violin plots showing distribution of expression of genes commonly upregulated in patients with translocations (y-axis), along with annotations of the cytogenetic alterations detected in samples by clinical iFISH assay (top).

After filtering cells using standard quality controls, we analyzed a total of 29,387 single CD138+ PCs (∼850 from MGUS, ∼8.4×10^3^ from SMM, ∼1.7×10^3^ from MM, and ∼9×10^3^ from NBM). Projecting cells onto a 2D Uniform Manifold Approximation and Projection (UMAP) plot, we observed that cells from our NBM samples (fresh or frozen) grouped together, while the majority of cells from patients with precursor conditions and overt MM formed separate groups of cells (Fig. 1b,c). The number of CD138+ cells analyzed per sample ranged from 40 to 3,414, with a median of 591.

Applying Leiden clustering^15^, we obtained 25 clusters of cells (Supplementary Table 3). Seven of these clusters represented healthy cells as determined by the majority of cells in these clusters coming from NBM samples and their overexpression of genes such as *CD27*. We merged these clusters into one “healthy” cluster (Fig. 1d). Of the remaining 18 clusters, 11 each consist almost exclusively of cells from a single sample, reflecting the fact that normal variation between healthy individuals was minor compared to disease-associated expression changes (Fig. 1e). With a few exceptions (Supplementary Note 1), the clusters that represented multiple samples grouped cells with shared disease biology: cluster 12 contained two sequential samples from the same patient, cluster 21 contained proliferating neoplastic cells from 15 patients across disease stages, and clusters 3 and 20 (together with cluster 24) represented all patients with a t(11;14) translocation.

Of note, our cohort included one patient, SMM-12, whose biopsy included two distinct subclones. Both subclones harbored a t(11;14) translocation, but only one acquired a CD20+ phenotype (Extended Data Fig. 1a), a MM phenotype occurring in up to 22% of patients^16^. This falls in line with previous studies that have shown CD20+ myeloma cells to be correlated with translocation t(11;14)^17^. Cells from the CD20-subclone clustered together with cells from other t(11;14) samples in cluster 20, while cells from the CD20+ subclone clustered separately in cluster 22, suggesting large expression changes associated with CD20+. Indeed, comparing gene expression in the CD20+ vs. CD20-subclones, we found 455 differentially expressed genes (DEG) (|log(fold change)| *>* log(1.5); false discovery rate q<0.1), with the top DEGs by q-value reflecting the B cell-like phenotype of these cells, including overexpression of *CD74*, *CD20* (also known as *MS4A1*), and HLA class II genes such as *HLA-DRA* and *HLA-DRB1* (Extended Data Fig. 1b; Supplementary Table 4).

To benchmark the performance of our experiment, we used interphase fluorescence in situ hybridization (iFISH) to identify large-scale structural genomic variants (Supplementary Table 1) and then inspected the expression levels of translocation target genes cyclin D1 (*CCND1*), MM SET domain (*MMSET/WHSC1*), fibroblast growth factor receptor 3 (*FGFR3*), MAF BZIP transcription factor (*MAF*), and MAF BZIP transcription factor B (*MAFB*), as well as cyclin D2 (*CCND2*) and Integrin Subunit Beta 7 (*ITGB7*), whose overexpression is also associated with translocations. MM cells from patients with iFISH-reported translocations exhibited overexpression of the respective target genes (Fig. 1f). In 4/7 samples whose iFISH results were inconclusive due to insufficient cell numbers, we were able to observe the overexpression of translocation partner genes or *CCND2* and *ITGB7*, indicating possible corresponding translocations. Our patient with sequential samples at SMM and after progression to MM confirms the ability of RNA-sequencing to capture the cytogenetic phenotype even prior to iFISH; while this patient’s iFISH results were inconclusive in the sample taken during SMM, we were able to detect overexpression of *ITGB7*, *MAFB*, and *CCND2* at the transcriptional level, suggesting a t(14;20) translocation, which was later confirmed by iFISH after the patient’s progression to MM.

### In silico dissection of normal and neoplastic cells within samples allows for characterization of disease even in samples with low tumor purity

One major benefit of analyzing single cell data from patients with precursor conditions is the ability to separate normal PCs from malignant and premalignant CD138+ cells (which we refer to inclusively as “neoplastic” cells). No individual marker genes can reliably distinguish these populations, but our full-transcriptome data enable aggregation of a neoplastic signal across many genes. To this end, we subclustered each individual sample based on its highly variable genes (but excluding genes located in immunoglobulin loci), and then examined patterns of immunoglobulin and MM driver gene expression in each subcluster in order to label the subcluster as containing normal or neoplastic cells (Extended Data Fig. 2). To complement and validate this method, we also developed a Bayesian hierarchical model for estimating the tumor purity of each individual sample based only on the distribution of immunoglobulin light chain expression. Comparing these results, we observe strong agreement between the two purity estimation methods (Fig. 2a).

**Figure 2.**
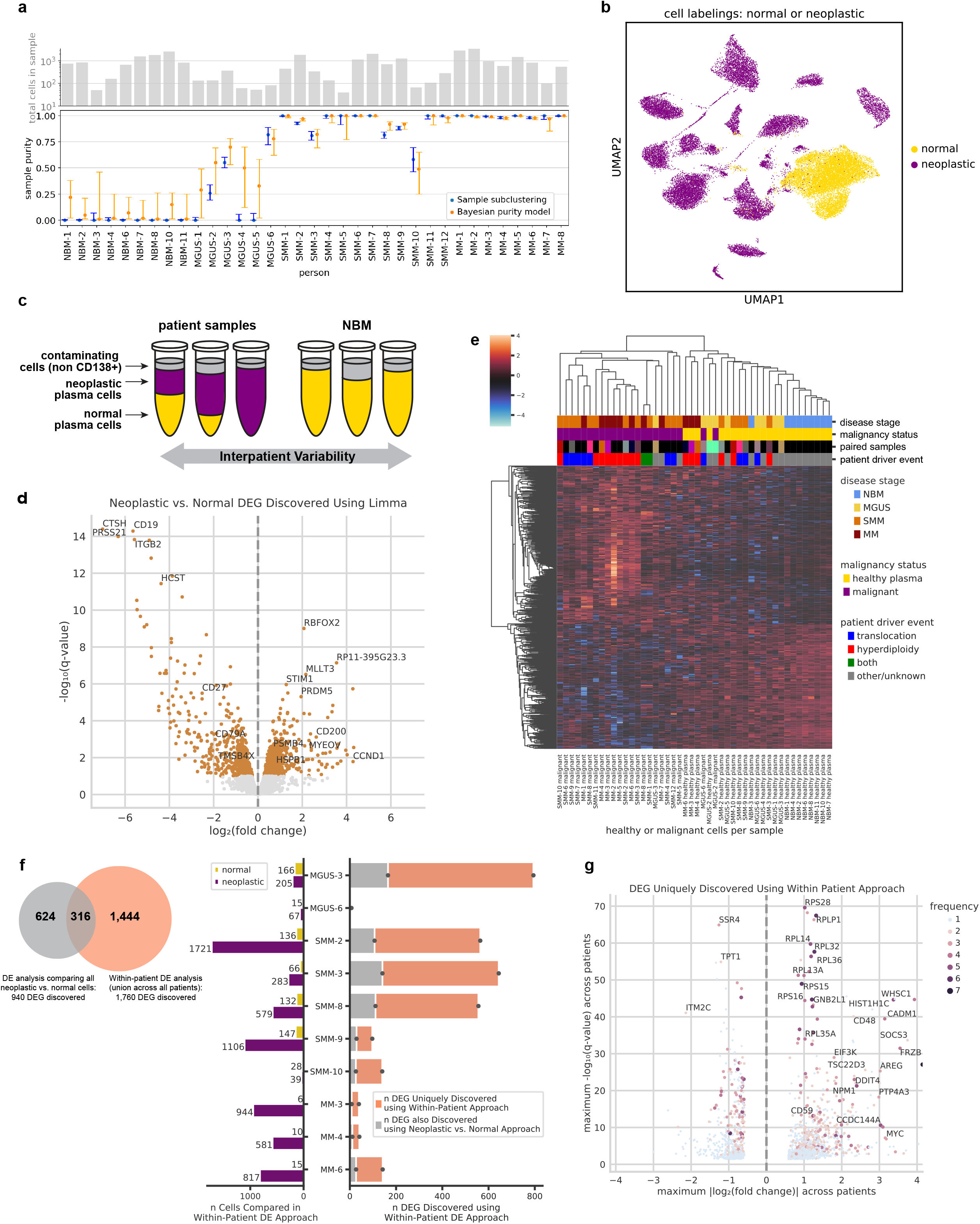
In silico dissection of transcriptional differences in healthy and malignant plasma cells within patient samples. **a,** The number of cells per sample (top), and the estimated purity of each sample with 95% confidence intervals (bottom). Sample purity was estimated using two orthogonal methods: subclustering of individual samples (blue) and our Bayesian hierarchical purity model (orange). Purity estimates for most samples have good agreement between these two methods. **b,** UMAP localization of individual cells labeled as normal or neoplastic. **c,** Cartoon schematic of our within-patient differential expression analysis. Whereas bulk studies would compare patient samples to NBM and be confounded by contaminating cell types and low sample purity, we isolate and characterize the neoplastic plasma cells in each patient. We run two DE analyses: In the first, we compare all neoplastic (purple) vs. all normal (yellow) cells using limma-voom. In our within-patient DE analysis, we instead compare patients’ neoplastic cells to their own healthy cells, controlling for inter-patient variability. Samples with 100% normal or neoplastic cells were excluded from the within-patient analysis. **d,** Volcano plot of limma-voom DE results for neoplastic vs. normal cell populations. Orange denotes genes with q-value<0.1. The 4 most significant up- and downregulated genes are annotated, as well as other selected genes. **e,** Pseudobulk expression of DEGs detected between neoplastic and normal pseudosamples using limma-voom (z-scored per gene). Each column represents the normal or neoplastic cells from a given sample. Color annotations denote disease stage (top), normal or neoplastic status (second), a mapping of paired columns coming from the same patient (third; matching colors denote that columns correspond to the same patient; black denotes that there was no paired sample), and whether IgH translocation or hyperdiploidy was detected in that sample by iFISH (bottom). The majority of normal and neoplastic pseudosamples cluster separately, regardless of disease stage or sample of origin. **f,** Quantification of DEGs uniquely discovered using within-patient DE. The venn diagram represents the overlap of DEGs found using the limma-voom and within-patient DE approaches, and the bar plot describes the number of DEGs found per-patient using within-patient DE (right side of bar plot). Since our power to detect DEGs using the within-patient method was affected by the number of cells (n) per patient, we show n neoplastic and normal cells per patient (left side of bar plot). **g,** Volcano plot of 1,760 DEGs uniquely discovered using our within-patient DE approach. The y-axis represents the maximum -log_10_(q-value) of the gene (across patients included in the within-patient analysis), and the x-axis represents the maximum log_2_(fold change). The color and size of a dot denote the number of patients for which that DEG was detected, with blue dots representing DEGs detected in just one sample.

Our purity results suggest that samples from patients with precursor conditions have a sizable fraction of healthy PCs, as expected. On average, MGUS samples contained 73% healthy cells and SMM samples contained 8%, compared to just 0.5% in MM. Furthermore, the variability of tumor purity values was also greater at early stages of disease. Whereas MM samples had consistently high tumor purity (range 0.98-1), we observed increasingly large variability in SMM (0.58-1) and MGUS (0-0.81), respectively. For our downstream analyses, we used each cell’s label from the subclustering approach in order to separate normal and neoplastic cells within each sample and characterized them independently. Our labels closely matched the Leiden clustering results (though not identically, highlighting the benefit of our curated labels), with 97% of cells we labeled as normal and <1% of cells we labeled as neoplastic belonging to the healthy Leiden cluster (Fig. 2b).

### Transcriptional differences between neoplastic and normal cells across patients

We performed a differential expression (DE) analysis comparing neoplastic and normal cells. To this end, we split samples into their neoplastic and normal populations, which we refer to as “pseudosamples.” We compared neoplastic pseudosamples to normal pseudosamples using limma-voom^18, 19^ and found 940 DEGs (|log(fold change)| *>* log(1.5); false discovery rate q<0.1).

In addition to genes known to be important for MM biology like *CCND1* (upregulated), *CD27* (downregulated) and*TMSB4X* (downregulated), we also found other strongly regulated genes whose connection to myeloma is less well characterized. The top 4 upregulated DEGs by q-value included *RBFOX2*, *MLLT3*, which is a proto-oncogene reported in acute myeloid leukemia^20^, *STIM1*, a transmembrane protein that mediates store-operated calcium entry and is of interest in multiple cancers^21–23^, and the long non-coding RNA *RP11-395G23.3*. The top 4 downregulated genes included *CTSH*, *CD19*, a B cell lineage marker gene which has been explored as a therapeutic target in MM despite its low expression^24, 25^, *PRSS21*, and *ITGB2*. We also observed upregulation of *PSMB4* and *HSPB1*, which are associated with the proteasome (Fig. 2d).

Unsupervised clustering of the pseudosamples based on their expression of DEGs showed good separation of neoplastic and normal samples, as expected, and also revealed that hyperdiploid patients exhibit especially high expression of the upregulated DEGs and tend to cluster together. Neoplastic samples do not cluster by disease stage, underscoring the fact that precursor cells from MGUS and SMM share a similar disease phenotype to myeloma cells. The two neoplastic samples that cluster together with normal cells came from MGUS patients with very low numbers of neoplastic cells detected (n=35 and 67 cells for MGUS-2 and MGUS-6, respectively). Thus we could not conclude that the MGUS phenotype is similar to that of normal cells, since the expression pattern of their pseudosamples is inherently noisy. Neoplastic cells from MGUS-3 (n=205), on the other hand, clustered together with other neoplastic samples (Fig. 2e).

Interrogating MSigDB hallmark genesets, we found that pathways related to E2F targets, Notch signaling, G2M checkpoints, interferon alpha response, and Wnt/beta-catenin signaling are differentially enriched in neoplastic samples compared to normal (t-test q<0.1; Extended Data Fig. 3a). When comparing pathway enrichment results between MGUS and normal samples, we found that the Wnt/beta-catenin pathway is already upregulated (t-test q=0.08). Individual upregulated genes from the Wnt/beta-catenin pathway include *DKK1*, *KAT2A*, and *TP53* (limma-voom neoplastic vs. normal q<0.025). Low sample size (n=3) for neoplastic MGUS samples may have hindered our power to discover other pathways that are already differentially enriched in MGUS vs. normal.

This DE analysis provides a general view of genes whose expression is consistently altered in disease, but it does not allow us to discover genes whose expression may be altered in just a small subset of patients in our cohort. Additionally, while normal cells are more similar to each other than neoplastic cells are, inter-patient differences still exist among them (Extended Data Fig. 4a,b). Thus, this analysis suffers from both high variance due to the small number of normal samples and confounding effects due to non-disease-related differences between individuals that contributed healthy bone marrow and tumor samples. We address these limitations with the following analysis.

### Within-patient neoplastic vs. normal cell comparisons highlight inter-patient heterogeneity and patient-specific disease characteristics

To account for the limitations of the DE analysis described above, we leveraged impure samples to perform a “within-patient” characterization of the disease. For each patient, we compared their neoplastic plasma cells to their own healthy plasma cells (Fig. 2c). This allowed us to specifically characterize the unique transcriptional profiles of individual tumors, which may not be shared across patients, without introducing the confounding effects that would arise from comparing tumor cells to normal cells from other healthy donors.

Of our eleven patients with both neoplastic and normal cell populations, ten had significant DEGs detected between these populations (|log(fold change)| *>* log(1.5); false discovery rate q<0.1). Overall, this method identified 1,760 DEGs, 1,444 of which were not found in the general neoplastic vs. normal DE analysis described above (Fig. 2f). We found DEGs that are unique to individual patients (1,323 genes) as well as genes recurrently affected across patients, such as *CD27* (upregulated in 8 patients), *CD79A* (upregulated in 7 patients), and *RPL25* (downregulated in 7 patients).

We next focus on genes that were not discovered in the general DE analysis described earlier (Fig. 2g). For example, within-patient DE enabled identification of significant upregulation of *FGFR3* and *WHSC1* in our patient with t(4;14). The general neoplastic vs. normal cell comparison was not powered to identify this upregulation, since the translocation only occurred in a single patient in our cohort. Additionally, we discovered upregulation of *GNB2L1* (also known as *RACK1*; up in SMM-2, SMM-3, MM-6), a known oncogene in other cancers^26, 27^ that has recently been reported to be upregulated in myeloma cell lines,^26, 27^ but not yet in clinical samples. Among upregulated genes, we also found the histone gene *HIST1H1C* (MGUS-3, SMM-2, SMM-3, SMM-8, MM-6), the cell surface markers *CD48* (MGUS-3, SMM-8) and *CD59* (MGUS-3, SMM-2, SMM-3, SMM-8, SMM-10), and the proto-oncogene *MYC* (MGUS-3, SMM-2, MM-6) (Fig. 2g; Extended Data Fig. 3b). We observed downregulation of *SSR4* (SMM-2, SMM-3, MM-3), associated with translocation of proteins across the endoplasmic reticulum, and *TPT1* (MGUS-3, SMM-8), a regulator of cell growth and proliferation. *ITM2C*, which has been reported for its expression on MM cells,^28, 29^ was upregulated in some samples (MGUS-3, SMM-2, SMM-3, SMM-8) but downregulated in others (MGUS-6, SMM-9). While higher expression of *ITM2C* has been reported in patients with t(4;14) vs. without^30^, we cannot conclude this from our data, as *ITM2C* was variably expressed in our 3 samples with t(4;14) (SMM-7, SMM-8, MM-1; Extended Data Fig. 3c). Ribosomal proteins such as *RPS28* (SMM-2, SMM-3, SMM-10, MM-3, MM-6), *RPLP1* (MGUS-3, SMM-2, SMM-3, SMM-10, MM-3, MM-4), *RPL14* (MGUS-3, SMM-2, SMM-3, SMM-10, MM-6), and others were recurrently upregulated, specifically in patients with hyperdiploidy. Although these ribosomal protein genes are upregulated in multiple samples, other samples have expression levels similar to those of NBM samples (Extended Data Fig. 3c), possibly explaining why they were only detected using within-patient DE.

### NMF discovers gene signatures that capture transcriptional programs

While our within-patient DE analysis allowed us to discover gene signatures in individual tumors, we next employed a method to discover gene signatures active in individual cells across our cohort, even if only in a small subset of cells in a tumor, and to characterize signature activity at the single cell level across disease stages.

Using our Bayesian-NMF method^31, 32^, we decomposed the gene expression profiles across all plasma cells in our cohort into 28 gene signatures, each of which represents a pattern of gene expression recurrently occurring across cells in our dataset (Supplementary Table 5). Because we were most interested in highlighting signatures associated with disease biology rather than patient-specific effects, we removed signatures that were only active in a single patient. Similarly, since our goal was to find groups of genes with shared activity patterns, we did not focus our downstream analyses on signatures that only represented the expression of a single gene. After removing these “patient-specific” and “single-gene” signatures, we retained 15 gene signatures and examined the top genes from each signature to identify its underlying biological mechanism (Fig. 3a; Table 1).

**Figure 3.**
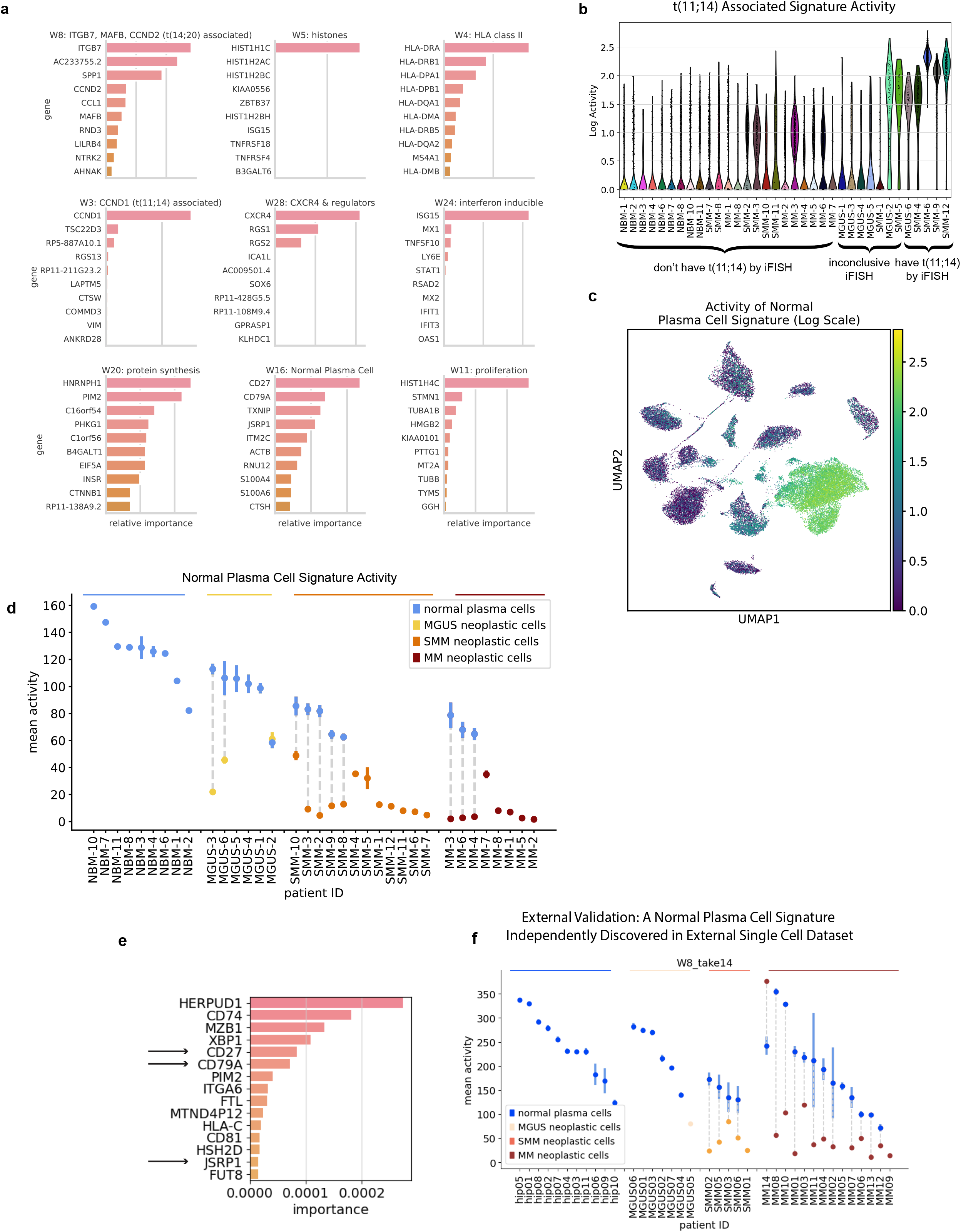
Bayesian non-negative matrix factorization uncovers gene signatures which capture myeloma cell biology across disease stages. **a,** Top genes for nine representative gene signatures. The importance score, plotted on the x-axis, is based on both the strength of the gene’s contribution to the signature and its specificity to the signature (see Methods). **b,** A signature with top contribution from *CCND1* is discovered and is most active in samples with t(11;14), as expected. **c,d** We discover a ‘normal plasma cell signature’ that is active in normal plasma cells across disease stages and downregulated in neoplastic cells from MM and precursor conditions. We visualize this signature’s activity on a UMAP plot (log scale) **(c)** and by showing its mean activity ± s.e.m. for the normal and neoplastic populations within each sample **(d)**. This signature is markedly downregulated in neoplastic cells as early as MGUS. **e,f** Validation on external dataset: our NMF algorithm run on external CD138+ single cell data from MGUS, SMM, MM and healthy donors independently discovers a gene signature similar to our normal plasma cell signature, with shared top genes *CD27*, *CD79A*, and *JSRP1* (e). After labeling cells in that dataset as normal or neoplastic, we discover that this signature follows the same pattern as in our data, with high activity in normal cells and a significant decrease in activity in neoplastic cells across disease stages. Mean activity ± s.e.m. of signature across normal and neoplastic portions of samples is shown **(f)**.

**Table 1.** : 15 gene expression signatures discovered using Bayesian NMF.

| Signature | Biological Description | Top Genes |
| --- | --- | --- |
| W3 | t(11;14) associated | CCND1, TSC22D3, RP5-887A10.1, RGS13 |
| W4 | HLA class II | HLA-DRA, HLA-DRB1, HLA-DPA1, HLA-DPB1 |
| W5 | histones | HIST1H1C, HIST1H2AC, HIST1H2BC, KIAA0556 |
| W8 | t(14;20) associated | ITGB7, AC233755.2, SPP1, CCND2 |
| W9 | extracellular signaling | LGALS1, VIM, ACTB, S100A6 |
| W11 | proliferation | HIST1H4C, STMN1, TUBA1B, HMGB2 |
| W16 | normal plasma cell | CD27, CD79A, TXNIP, JSRP1 |
| W20 | protein synthesis | HNRNPH1, PIM2, C16orf54, PHKG1 |
| W24 | interferon inducible | ISG15, MX1, TNFSF10, LY6E |
| W28 | CXCR4 & regulators | CXCR4, RGS1, RGS2, ICA1L |
| W1 | unknown | JUNB, ZFP36, NFKBIA, IER2 |
| W6 | unknown | DUSP4, GADD45A, BTG2, LAMP5 |
| W14 | unknown | KLF6, TSC22D3, ANKRD28, KLF2 |
| W26 | unknown | HLA-A, ITM2C, PRR15, ACTB |
| W27 | unknown | NEAT1, DDX17, ANKRD12, FOXO3 |

A number of the NMF signatures represent subtypes of myeloma that have previously been reported; for example, we find a signature of proliferation similar to that reported by Zhan et al.^33^ and Broyl et al.^34^, a *CCND1*-related signature which is differentially active in samples with t(11;14) (Fig. 3b), and a signature composed of *MAFB, CCND2,* and *ITGB7*, which is active in the samples with t(14;20).

We additionally discovered signatures that elucidate less well-characterized disease biology that is common to multiple samples in our cohort. For example, we found a signature with top genes *CXCR4*, which plays a role in normal plasma cell development and has also been implicated in MM progression^35, 36^, and *RGS1* and *RGS2*, which are regulators of G Protein signaling that may regulate the CXCR4-CXCL12 axis^37^. We additionally found signatures that represent the activity of histone genes, interferon (IFN)-inducible genes, and genes involved in protein synthesis, among others.

### Gene signature activity correlates with disease stage and microenvironment

For each gene signature, we tested whether its activity level varied between malignant and normal cell populations or with disease stage, and discovered that three signatures had significantly different activity levels across disease stages (Kruskal-Wallis *p <* 0.05 and Dunn’s *q <* 0.1). As expected, we found that the t(11;14)-related signature is differentially active in the neoplastic cell population from samples with the corresponding translocation compared to NBM cells. We found two additional signatures whose activity correlated with disease stage, described in detail below.

### Neoplastic cells across disease stages share universal downregulation of gene signature seen in normal PCs

We discovered a “normal plasma cell signature” that is downregulated in neoplastic cells at all stages of disease (q<0.1; the difference between MGUS malignant cells and normal plasma cells did not reach significance, likely due to low number of MGUS cases with sufficient neoplastic cells, but the trend was still observed). This signature robustly characterizes the normal bone marrow plasma cells in our data set (Fig. 3c,d), and highlights genes that are downregulated only in the neoplastic cell portions of samples at all disease stages. The top genes in this signature include *CD27* and *CD79A*, which are associated with the B cell lineage, and *JSRP1*, *CTSH*, *HCST*, and *RNU12*, genes as of yet unreported to be involved in plasma and MM cell biology (Fig. 3a; Extended Data Fig. 5a). Other canonical B cell markers were not expressed (*CD20*, *BCL6, PAX5,* and *E2F1)* on healthy PCs suggesting this effect is not due to B cell contamination (Extended Data Fig. 6a,b). Given the low tumor purity during early precursor conditions, this phenotype would be obscured at early disease stages in bulk samples; analysis at a single cell resolution, however, reveals that this healthy plasma phenotype is significantly downregulated in malignant cells as early as the MGUS stage (Fig. 3d). Indeed, for our patient with serial samples at the SMM and MM stages (SMM-1 and MM-8), we find similarly low levels of signature activity at these two timepoints, underscoring the fact that this phenotype is lost at early stages of malignancy and remains low as the disease progresses (Fig. 3d). Interestingly, the activity of this signature in normal cells also trends downward with increasing disease stage (Jonckheere-Terpstra test p=1.3×10^-5^).

The individual top genes on this signature are also downregulated in a given patient’s neoplastic cells compared to its own healthy cells (Extended Data Fig. 5c). The notable exception is our CD20+ sample: while *CD27* is upregulated in neoplastic cells vs. normal cells in this sample, the NMF normal plasma cell signature nonetheless has low overall activity (Fig. 3d), demonstrating the universal loss of this signature across tumors with different phenotypes.

### Validation of normal plasma cell signature in independent datasets

To validate our findings, we ran the Bayesian NMF algorithm on single cell data from Ledergor et al.^12^ and recovered a similar signature with top genes *CD27*, *CD79A*, and *JSRP1*. This signature, too, is strongly downregulated in neoplastic cells as early as MGUS (Fig. 3e,f). As additional validation in bulk data, we scored bulk RNA sequencing from newly diagnosed MM samples in the Multiple Myeloma Research Foundation’s (MMRF) CoMMpass dataset for their expression of this signature, as well as for tumor purity. We found a significant negative correlation between signature expression and tumor purity, further supporting this signature as a marker of normal plasma cells (Extended Data Fig. 5b).

### Interferon-inducible signature upregulated in tumor and microenvironment

We discovered a signature enriched for IFN-inducible genes, such as ISG15 ubiquitin like modifier (*ISG15*), MX Dynamin Like GTPase 1 (*MX1*), and Interferon Induced Protein With Tetratricopeptide Repeats 1 and 3 (*IFIT1* and *IFIT3*) (Fig. 3a). Notably, this signature is significantly upregulated in both normal and malignant populations from overt MM patients compared to NBM (*q* = 0.012 and *q* = 0.0008, respectively; Fig. 4b). This upregulation is specific to malignant disease (signature is not significantly upregulated in precursor conditions; one patient, SMM-11, is an outlier with very high activity). Further, similar IFN-inducible signatures were discovered when running Bayesian NMF on T cell and CD14+ monocytes from the microenvironments of these patients’ tumors^14^, and these signatures are active in the same patients that have high IFN-inducible gene expression in their CD138+ cells (Fig. 4a). T cell and monocyte markers were not expressed in CD138+ cells, suggesting this correlation is not due to cell type contamination (Extended Data Fig. 6c).

**Figure 4.**
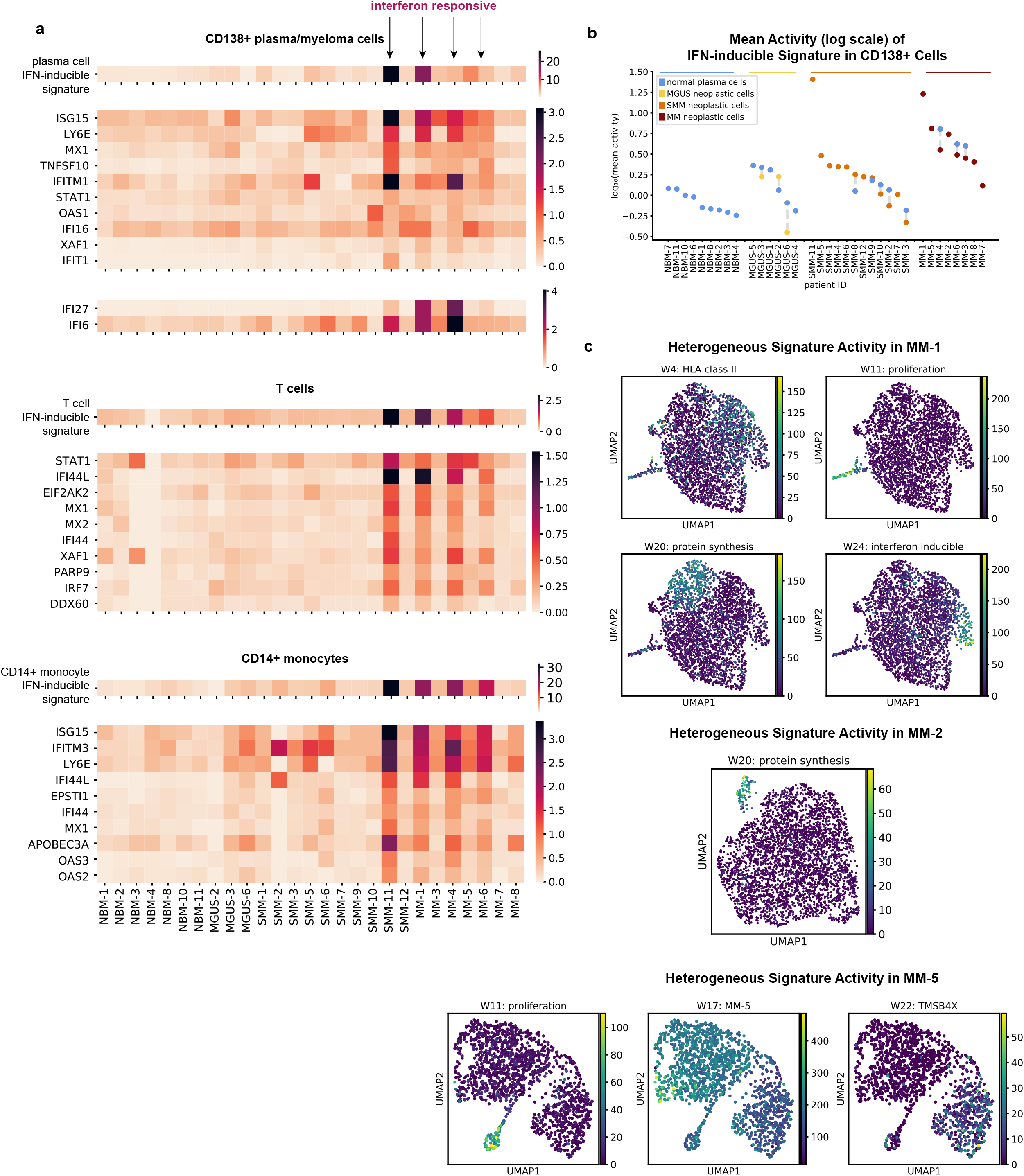
Correlation of IFN-Inducible Signature in CD138+ and Microenvironment Cells and Intratumor Heterogeneity of Signature Activities. **a,** Mean activity per sample of IFN-inducible signature discovered in CD138+ cells (top), T cells (middle) and CD14+ monocytes (bottom). Mean expression levels for the ten genes with the highest values in the W matrix for each signature are also shown. Expression of additional interferon-inducible genes *IFI27* and *IFI6* is shown for CD138+ samples (see Supplementary Note 2). CD138+ samples from patients were limited to neoplastic cells before calculating means. **b,** Mean activity ± s.e.m. of plasma cell IFN-inducible signature across normal and neoplastic plasma cell populations. Both normal and neoplastic plasma cells exhibit significantly increased activity of the interferon-inducible signature in MM vs. normal bone marrow (q=0.012 and q=0.0008, resp.). **c,** Subpopulations within patient tumors heterogeneously express gene signatures. Cells from a given sample were projected onto a UMAP plot based on expression of highly variable genes, and colored by signature activity. In MM-1, disjoint subsets of cells express the HLA class II, IFN-inducible, proliferation, and protein synthesis signatures. Heterogeneously expressed signatures for MM-2 and MM-5 are also shown.

### Tumors contain transcriptionally heterogeneous cell subpopulations

The NMF approach to signature discovery allows us to find groups of genes with shared activity in single cells, and thus not only to examine how signature activity varies between samples and disease stages, but also between subpopulations of cells within a single sample. Indeed, we find that tumors are heterogeneous, with subsets of cells expressing distinct gene signatures. For example, in patient MM-1, disjoint subsets of cells express the HLA class II, IFN-inducible, proliferation, and protein synthesis signatures; this is discernible on a UMAP plot of patient MM-1’s cells colored by the activity level of these signatures (Fig. 4c) or the top genes from these signatures (Extended Data Fig. 7a). Similarly, patient MM-2 contains only a subset of cells with high expression of the protein synthesis signature, and patient MM-5 contains a subset of proliferating cells, as well as varying activity of signature 17 (an MM-5 patient-specific signature with top genes *EIF3L*, *MYC*, *RPL36A*, and *JUNB*) and signature 22 (a single-gene signature representing the expression of *TMSB4X*) (Fig. 4c; Extended Data Fig. 7b,c).

## Discussion

Early identification of precursor MM conditions and those patients that are at risk of progressing to overt MM is critical as it allows for early therapeutic interventions in these patients. However, the current risk criteria used to identify high risk precursor patients who would most benefit from treatment are only based on clinical parameters such as M spike, light chains or percent tumor burden. Therefore, elucidation of the molecular transformation that occurs at early tumorigenesis and later at high risk SMM before disease progression is critical for developing informed criteria for patients who would benefit from early intervention and targets that may be exploited for therapeutics.

Here, we leveraged single-cell RNA sequencing to overcome the challenges of characterizing the transcriptomics of these low burden precursor states. While some patients at early stages of disease had low disease burden, such that their driving cytogenetic translocations could not yet be detected by iFISH in the clinic, we demonstrate that RNAseq is sensitive enough to already reveal their underlying cytogenetic changes. This result highlights the possibility of using RNAseq to detect phenotypic changes in patients’ bone marrow plasma cells earlier than methods currently used in the clinic.

Precise labeling of normal and neoplastic cells in each sample revealed low tumor purity in samples from earlier disease stages, even when subsetting to only CD138+ cells. This suggests that conclusions drawn from bulk studies of precursor conditions are likely influenced by heavy contamination by normal plasma cells. For example, Chng et al.^11^ conclude that their “MYC activation signature” is upregulated in a subset of myelomas, but not in MGUS. While it is possible that MYC activation really did not occur in their MGUS samples, we do find *MYC* and some of their MYC activation signature genes to be significantly upregulated in two precursor patients in our cohort (MGUS-3 and SMM-2), as well as in two MM patients (MM-5 and MM-6). The upregulation of *MYC* in MGUS is clinically relevant, as it is associated with tumor aggression and poor clinical outcomes^38^. Both the low tumor purity in MGUS and the potential rareness of this phenotype among MGUS patients would have made this difficult to discover without single cell data, the ability to distinguish normal vs. neoplastic cells, and our within-patient DE analysis.

Through our isolation of neoplastic cells, we found that these early pre-cursor conditions already exhibit transcriptomic alterations seen in overt MM. As one important example, we identified a signature present in normal plasma cells but uniformly lost at all stages of MM progression. *CD27*, one of the top genes of this signature, has been previously discussed in MM literature, but has been reported to have variable expression in myeloma cells, increased expression in MGUS, and a correlation with prognosis^39–41^. Our data shows significant downregulation of *CD27* compared to normal plasma cells as early as the MGUS stage (Supplementary Table 4; Extended Data Fig. 5c). Although we observe a trend of decreased *CD27* expression in MM compared to SMM (Extended Data Fig. 5c), it raises the question of the extent to which previous results were confounded by increasing tumor purity as the disease progresses. This would need to be tested in a larger cohort with single cell data.

In addition to *CD27*, neoplastic plasma cells also had lower expression of another mature B cell marker, *CD79A*, as well as decreased enrichment of immune pathways, such as complement pathway (including decreased expression of the complement receptor 2, *CR2*, also known as *CD21*). Similarly, other studies found absence or low levels of B cell surface markers CD19, CD27, and CD45 on neoplastic cells compared to normal plasma cells^39, 42^. Our study extends the characterization of matched neoplastic and normal plasma cells to the whole transcriptome. It also supports the hypothesis that the loss of B cell immune functionality, as assessed by gene expression programs and cell surface protein expression, is an early step in the generation of tumor plasma cells.

When probing pathway level transcriptional changes in neoplastic cells, we found aberrant expression of Wnt pathway members including overexpression of *DKK1* in neoplastic cells of precursor myeloma (Extended Data Fig. 3c). DKK1 is secreted by myeloma cells and is associated with the presence of osteolytic lesions through inhibition of osteoblast differentiation^43, 44^. Given that many cases of MGUS also have osteoporosis and osteopenia, this may indicate that Wnt dysregulation and *DKK1* overexpression are associated with early osteopenia in those patients and are potentially predictors of the development of osteolytic lesions.

When investigating connections between the phenotype of PCs and dynamics in the tumor immune microenvironment, we discovered that patients who exhibit upregulation of an IFN-inducible signature in their tumor cells exhibit this same phenotype in their normal bone marrow PCs, as well as in T cells and monocytes in their tumor microenvironment. This suggests that interferon signaling in myeloma cells, which has been reported previously^45^, may be a response to a common stimulus in the microenvironment that affects multiple cell types, including normal plasma cells. Further work is needed to investigate the potential common mechanisms driving the upregulation of interferon signaling across these cell types.

The single cell resolution of our data provides insight into the heterogeneity of signatures within patient samples. For example, while the proliferation signature has previously been reported in bulk studies as characterizing a distinct subset of patients^33^, our data reveal that in fact, only a subset of cells from any given patient exhibit a proliferative signature. Additionally, these proliferating cells may be found in patients that harbor the driver mutations previously used to characterize patients that belong to the non-proliferative subtypes.

Finally, our within-patient DE analysis points to potential therapeutic targets that are present in subsets of patients not only in MM, but also in earlier disease stages, which can be selected for functional validation. We found that select patients exhibit upregulation of genes associated with the proteasome, which may correlate with sensitivity to bortezomib and antitumor immune response^46^. While certain proteasome genes were identified in our general neoplastic vs. normal DE analysis (*PSMB4*, *HSPB1*), we discovered upregulation of additional proteasome genes in select patients using our within-patient DE approach (e.g. *PSMA4*, *PSMD14*, *PSMD11*, *PSMC6*, and *PSMA1* in MGUS-3 and SMM-8), painting a much fuller picture of proteasome-related gene enrichment in those samples. Additionally, this approach allowed us to detect that *CD59*, a complement inhibitor whose expression has been associated with resistance to daratumumab in myeloma^47^ and to anti-CD20 therapies in B cell malignancies^48^, was significantly upregulated in five patients, including those with precursor conditions. In a similar vein, we discovered upregulation of *CD48*, which has been nominated as a drug target in MM^49^, in neoplastic cells from MGUS and SMM. Our identification of patient-specific transcriptional changes as early as MGUS paves the way for future work exploring personalized treatment approaches prior to malignant disease.

In summary, our work used single-cell RNA sequencing to overcome the low fraction of neoplastic plasma cells in early precursor disease and uncovered previously unidentified transcriptional changes that occured at the precursor stage and could not be described in prior bulk sequencing studies. We identified previously-unappreciated commonalities between MGUS and overt MM, such as the loss of our novel normal plasma cell signature. Elucidating patient-specific transcriptional changes, as our within-patient DE analysis does, may allow for precision medicine approaches to treating MM and potentially intercepting precursor conditions before progression.

## Methods

### Patient samples and cell preparation

Primary peripheral blood cells or BM samples from patients with MGUS, SMM or MM, as well as healthy donors were collected at the Dana-Farber Cancer Institute. These studies were approved by the Dana-Farber Cancer Institute Institutional Review Board. Informed consent was obtained from all patients and healthy volunteers in accordance with the Declaration of Helsinki protocol (fifth revision from 2000 with Clarifications of Articles 29, 30 (20022004), and the most recent iteration from 2013). MGUS and SMM patient samples were collected for a clinical trial, clinicaltrial.gov identifier NCT02269592. CD138+ BM cell fractions were isolated using magnetic-activated cell sorting technology (Miltenyi Biotec). Selected cells were either viably cryopreserved in dimethylsulfoxide at a final concentration of 10% or used immediately for scRNA-seq.

### Sequencing library construction using the 10x Genomics platform

Frozen BM cells were rapidly thawed, washed, counted and resuspended in PBS and 0.04% bovine serum albumin to a final concentration of 1,000 cells per µl. The Chromium Controller (10x Genomics) was used for parallel sample partitioning and molecular barcoding. To generate a single-cell Gel Bead in EMulsion, cellular suspensions were loaded on a Single Cell 3’ chip together with the Single Cell 3’ Gel Beads, according to the manufacturer’s instructions (10x Genomics). scRNA-seq libraries were prepared using the Chromium Single Cell 3’ Library Kit v.2 (10x Genomics). Fourteen cycles were used for the total complementary DNA amplification reaction and for the total sample index PCR. Generated libraries were combined according to Illumina specifications and paired-end sequenced on HiSeq 2500/4000 platforms with standard Illumina sequencing primers for both sequencing and index reads; 100 cycles were used to sequence Read1 and Read2.

### Preprocessing of scRNA-seq data

Sample demultiplexing, barcode processing, alignment to the human genome (hg38) and single-cell 3’ gene counting was performed using the Cell Ranger Single-Cell Software Suite v.2.0.1. Cells called by Cell Ranger were further filtered to those with <15% mitochondrial expression, >200 genes covered, <50,000 total unique molecular identifiers (UMIs), and <4,000 total genes detected. Log-normalized expression values were calculated as:

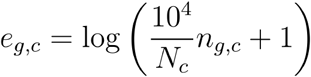

for a cell *c* with *N_c_* total UMIs from genes (excluding genes that accounted for >20% of UMIs in any cell), with *n_g,c_* UMIs mapping to gene *g*. Except where noted, “expression” refers to log-normalized expression.

### Gene selection

For downstream analyses (PCA, UMAP, Leiden clustering, differential expression, NMF), we removed genes located in IGH, IGL, or IGK loci (based on the GRCh38 reference), since these are expected to be upregulated and clonally expressed in neoplastic cells, dominating other transcriptional disease signatures of interest. Sex genes XIST and RPS4Y1 were also removed prior to PCA, UMAP, clustering, and NMF, so as not to separate samples based on the sex of the patient but rather based on disease biology. Highly variable genes were selected based on log-normalized expression data using the highly_variable_genes function in Scanpy version 1.7.1^50^ with default parameters and max_mean=4, except where indicated otherwise.

### Removing non-CD138+ cell populations

To remove cells incorrectly sorted during bead selection, we first performed coarse clustering of all cells sorted as CD138+. We centered and scaled the data, clipping the resulting values to a maximum of 10, calculated highly variable genes, projected the expression of highly variable genes onto its first 14 principal components, and computed Leiden clusters (resolution=1.5), all using the Scanpy package with default parameters except as specified. We chose this high resolution for clustering as our goal was to find and remove even small clusters of contaminating non-CD138+ cells. Using expression of known cell type markers, we identified and removed clusters of cells containing non-CD138+ immune cells, red blood cells, and cells from the extracellular matrix.

### Leiden clustering of CD138+ cells

After removing contaminating cell types, we reprocessed our data prior to downstream analyses. Despite removing clusters of red blood cells, we still detected ambient contamination of hemoglobin genes in some samples, and thus we regressed out a “hemoglobin score,” computed as the mean of log-normalized expression of hemoglobin genes. We then recomputed highly variable genes, re-centered and scaled the data, clipping the resulting values to a maximum of 10, projected the expression of highly variable genes onto its first 14 principal components, and computed Leiden clusters (resolution=1.5), all using the Scanpy package with default parameters except as specified. Here, we again chose a high resolution for clustering in order to detect even small clusters of unique cell types. We merged seven clusters which were all determined to contain healthy cells based on the majority of cells in these clusters coming from NBM samples, their overexpression of genes such as *CD27*, and their co-localization on a 2D UMAP embedding plot.

### Bayesian Model for Sample Purity Estimation

We developed the following hierarchical Bayesian model to automatically estimate ρ, the purity of a sample, based on the ratio of kappa:lambda immunoglobulin gene expression in that sample.

We assume the following generative model:

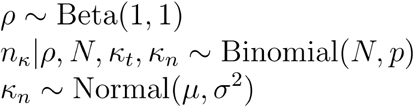

Where *ρ* is sample purity (drawn from a uniform distribution), *n_κ_* is the total number of kappa cells in a sample, *N* is the total number of cells in a sample, *κ_t_* is the proportion kappa cells among neoplastic cells in a sample (either 0 or 1 due to clonality), and *κ_n_* is the proportion kappa cells among normal cells in a sample. Cells were defined as kappa or lambda based on whether they have higher expression of *IGKC* or *IGLC2*, respectively. *µ* and ^2^ are empirically estimated from the NBM samples in our cohort, and *p* is the expected proportion of kappa cells, given *κ_t_*, *κ_n_* and

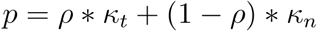

We calculate the joint probability of the light chain type of the tumor and the tumor’s purity, given the number of kappa cells and the total number of cells in the sample, and then marginalize to find the probability of tumor purity values:

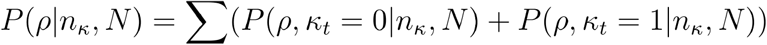

Assuming that *ρ* and *κ_t_* are independent of each other and of *N*, we can calculate the desired posterior joint probability using the following expression:

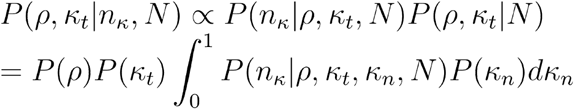

We report the mode of *P*(*ρ*) as the purity estimate, along with 95% confidence intervals.

A python implementation of this purity model will be made available on GitHub prior to publication.

### Sample subclustering approach to labeling normal and neoplastic CD138+ cells

For each sample, we performed a cluster analysis of only the cells in that sample. More specifically, we calculated variable genes based on log normalized expression using Scanpy’s highly_variable_genes function with parameter min_disp=0.6, centered and scaled the data, clipping the resulting values to a maximum of 10, and ran PCA and Leiden clustering (determining the number of PCs to input to Leiden clustering based on an elbow plot). We manually inspected the resulting clusters for each sample to determine whether each cluster contained healthy or neoplastic cells. This determination was based on whether the cluster uniquely expressed the clonal immunoglobulin for that tumor (since immunoglobulins were removed from the highly variable gene list, they did not influence the clustering results), as well as each cluster’s expression of certain oncogenes, such as *CCND1* for t(11;14) tumors. All cells were labeled in this way, except for 20 cells from sample MGUS-2 that were characterized by low expression of *MALAT1* and were not obviously similar to the healthy or neoplastic cells from that sample, and thus were not classified and were excluded from downstream analyses. See Extended Data Fig. 2 for an example of this method applied to a sample.

In addition to labeling each individual cell as normal or neoplastic, this approach allowed us to determine a tumor purity for each sample, i.e. the fraction of cells labeled neoplastic. To calculate confidence intervals on this purity estimate, we assumed that our observed data was generated as *n* ∼ Binomial(*N,p*), where *N* is the total sequenced cells in a sample, *n* of which we labeled as neoplastic, and *p* are the proportion of neoplastic cells in the patient sample (not just the ones we sequenced). We further assume a uniform prior on *p* (*p* ∼ Beta(1,1)), thus the posterior distribution on *p* is *p*|*n,N* ∼ Beta(*n* + 1*,N* − *n* + 1), by conjugacy of the Beta and Binomial distributions. We derived 95% confidence intervals on each sample’s purity estimate based on the inverse cdf of its beta-distributed posterior.

### Neoplastic vs. normal differential expression testing with limma

DEGs between neoplastic and normal cells were derived using limma version 3.42.2 with voom transformation^18, 19, 51^. Samples were split into their neoplastic and normal populations, and we refer to each of these as a “pseudosample.” Counts across cells in a pseudosample were summed and used as input to the limma-voom pipeline. Immunoglobulin genes and genes with counts per million (CPM) <5 in all samples or expressed in <5% of both neoplastic and normal cells were removed prior to analysis, resulting in normalization and DE testing of 6,521 genes. Pseudosamples were normalized using the trimmed mean of M values (TMM) method^52^ and fold changes were calculated as implemented in limma. We controlled for age and sex. Age information was missing for one NBM sample, and we filled it using mean imputation based on the ages of the other NBM samples. DEGs were those with a Benjamini-Hochberg FDR<0.1 and |log fold change| > log(1.5).

### Within-patient differential expression testing

For each patient with both neoplastic and normal cells detected, we calculated DEGs between their neoplastic and normal cell populations using a Wilcoxon rank sum test, correcting for multiple hypothesis testing across genes tested for each patient. We calculated fold changes as implemented in Scanpy, but replacing their offset term of 1⨉10^-9^ with half of the minimum (non-zero) log-normalized expression value in our data (0.126), to avoid inflating fold changes.

Specifically, fold change was calculated as the ratio of

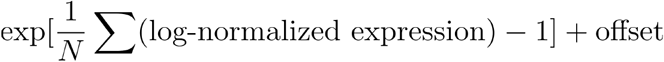

in each group, where *N* denotes the number of cells in the group. Differentially expressed genes were those with a Benjamini-Hochberg FDR<0.1 and |log fold change| > log(1.5).

For visualizing DEG uniquely found using this within-patient DE approach (Fig. 2g), we first limited DEG to those not found using limma. Then, for each gene, we calculated a maximum log_2_(q-value) as the maximum (BH-corrected) q-value reported across patients, multiplied by the number of patients with DEG (10, in our data) to further correct for multiple hypothesis testing across patients. This value was calculated separately for upregulated and downregulated instances of DEG, where applicable.

### Bayesian nonnegative matrix factorization (NMF)-derived gene expression signatures

We defined gene expression signatures using our SignatureAnalyzer-GPU tool^32^ (https://github.com/broadinstitute/SignatureAnalyzer-GPU), which implements a previously described Bayesian NMF algorithm^31^. This method approximates the expression profile of each cell (represented as a column in the input matrix, **V**) as an additive combination of latent expression programs (each column in the **W**-matrix output), each with an associated weight or ‘activity’ in each cell given by the **H**-matrix:

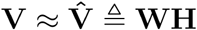

This Bayesian variant of NMF encourages sparse interpretable solutions by imposing an exponential prior distribution on the weights of the **W**- and **H**-matrices and allows automatic discovery of the number of signatures (*K*) required to explain the data.

After signature discovery, the columns of **W** were normalized to a sum of 1 and all the weight was shifted into the **H**-matrix:

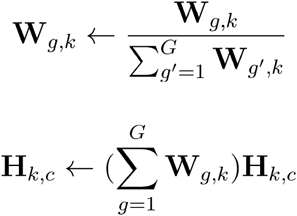

for gene *g* (out of *G* total genes), signature *k* and cell *c*.

Our input data was UMI counts for 3,883 highly variable genes, which were determined using Scanpy’s highly_variable_genes function with min_disp=0.2, which we set to be lower than the default value so as to include genes which may have relevance to plasma and myeloma cell biology despite a modest dispersion value. In addition to other default settings, we use a Poisson objective with an L1 prior on **W** and an L2 prior on **H**, set the initial *K* to 50, the maximum number of iterations to 7,000, and the tolerance to 1⨉10^-5^. We held out 20% of cells as a validation set. Since the Bayesian NMF algorithm finds a local minimum each time it is run, we ran the algorithm 100 times on our data in order to choose an optimal solution. Over 100 runs, the algorithm returned solutions with *K* between 24 and 30 with a mode of 28, and we chose the set of signatures with the lowest beta divergence over the validation set from among the solutions with *K* = 28. Before analyzing the signature results, we normalize each column in **H** by that cell’s total counts.

A signature was classified as “patient-specific” if its mean activity across cells in any one patient is >4 standard deviations higher than in all other patients. Otherwise, a signature is classified as “single-gene” if the weight of its most highly weighted gene based on the **W** matrix is ≥ 0.5 more than the weight of its next highest weighted gene. If a signature doesn’t meet either of these criteria, we describe it according to its top genes, where signature genes are ranked by their weight in **W** multiplied by their specificity to that signature, with specificity *S* defined as:

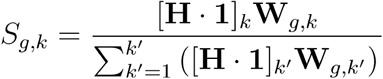

Signatures significantly altered between disease states were identified by calculating the mean signature activity for the neoplastic and normal cell populations in each sample, respectively, and performing a Kruskal-Wallis one-way analysis of variance and Dunn’s multiple comparison test with Bonferroni correction to detect differences in mean activities between the following groups: NBM, normal MGUS, normal SMM, normal MM, neoplastic MGUS, neoplastic SMM, and neoplastic MM. Comparisons with family-wise error rate < 0.1 were considered significant.

We additionally ran Bayesian NMF on an external single cell dataset^12^ using the same methods as above. We first limited the external data to cell types and disease stages which are present in our data, retaining only bone marrow PCs derived from healthy donors and patients with MGUS, SMM or MM, and limited the input features to hypervariable genes across these cells (4,669 genes).

### Signature scoring in the MMRF dataset

To calculate a signature score for each sample in the MMRF bulk RNA-sequencing dataset, we first used the MMRF’s publicly available counts data as input to calculate log-normalized transcripts per million (tpm) using the DESeq2 method for size factors.^53^ Then, for top signature genes (for our normal plasma cell signature, these included *CD27*, *CD79A*, *RNU12*, *JSRP1*, *SAT1*, *CTSH*, and *HCST*), we z-scored log-tpm expression of each gene across samples and calculated the mean of z-scored gene expression values.

### Pseudobulking Procedure

To pseudobulk samples, we summed the gene counts across cells, calculated the total gene counts in the sample (ignoring genes that accounted for >5% of counts), divided the summed count vector by the total gene counts, and multiplied by one million.

### Single Sample GSEA (ssGSEA)

Samples were split into their neoplastic and normal populations, and we refer to each of these as a “pseudosample.” We calculated the pseudobulk expression for each pseudosample and input this to the ssGSEA module available on the GenePattern platform^54, 55^ to calculate enrichment scores for the hallmark gene sets provided by the Molecular Signature Database (MSigDB)^56^. We removed pseudosamples comprised of <20 cells from downstream analysis of ssGSEA results, due to the high variance inherent in their gene expression. Differential pathway activity between two groups of pseudosamples was calculated using a t-test, and pathways with BH FDR < 0.1 were reported.

### Statistical analysis

Kruskal-Wallis one-way analysis of variance and Dunn’s multiple comparison test with Bonferonni correction were used when three or more independent groups were compared. When comparing two independent groups, all parametric tests were two-tailed, and the Benjamini-Hochberg (BH) method was used to correct for multiple hypothesis testing where appropriate. *P <* 0.05 or *q <* 0.1 (in cases of multiple hypothesis correction) were considered statistically significant. Error bars plotted on visualizations of mean signature activity or gene expression in a sample represent the standard error of the mean, and were calculated as the standard deviation of the means of 10,000 bootstrapped versions of that sample.

## Data Availability

All data analyzed in the present study will become publicly available when the study is published in a peer-reviewed journal, and are currently available upon reasonable request to the authors.

https://research.themmrf.org/

## Supplementary Information

### Supplementary Note 1

While the cells in cluster 17 passed our QC pipeline and were therefore included in the analysis, cells in this cluster are characterized by extremely low expression of *MALAT1*, as well as a low number of genes detected overall (median number of genes detected is lowest in cluster 17 compared to all other clusters), likely signifying a cell quality issue. Clusters 11 and 19 each grouped cells with no specific translocation and a relatively low number of neoplastic cells sequenced.

### Supplementary Note 2

Two gene signatures discovered in the CD138+ cells involved interferon-inducible genes: W24 (the plasma cell “IFN inducible” signature), whose activity is shown in the topmost heatmap in Fig. 4a, and W7, a patient-specific signature which involves many varied genes highly expressed in sample MM-4, including IFI27 and IFI6. Because W7 also involves non-interferon related genes, we do not show it in the main figure, but MM-4’s high expression of interferon-inducible genes is captured by that signature (mean activity of W7 in MM-4 = 276) instead of W24.

### Supplementary Tables

**Supplementary Table 1. Patient data and clinical information.** List of clinical measurements for samples used for single-cell RNA sequencing. **Supplementary Table 2. Sample information.** Quality metrics for scRNAseq samples, including whether the sample was fresh or frozen, n cells retained after removing low quality cells and non-CD138+ cells (QC), and median UMI post-QC. **Supplementary Table 3. Cell level information.** Sample of origin, Leiden clustering assignment, normal/neoplastic label, n genes detected, fraction mitochondrial reads, and n UMI detected per cell, for cells retained post-QC. **Supplementary Table 4. DEGs between CD20+ and CD20- subclones in SMM-12.** List of DE testing results for cells in SMM-12’s CD20+ vs. CD20- subclones. A Wilcoxon rank sum test with BH correction was used, and fold changes were calculated as described in methods (“Within-patient differential expression testing”). **Supplementary Table 5. Full list of NMF signatures.** List of top genes and descriptions for all 28 signatures discovered using Bayesian NMF.

### Data Availability

The scRNA-seq data that support the findings of this study are deposited with the NCBI Gene Expression Omnibus (GEO), accession number GSE193531, and will be made available upon publication. Updates to existing DbGaP repository (phs001323.v2.p1) are underway to reflect additional samples included in this analysis; a new version of the existing dbGaP repository will be available before publication and the accession number will be updated accordingly. All other data supporting the findings of this study will be available from the corresponding author at the time of publication upon reasonable request.

### Code Availability

The single-cell RNA data was processed using cellranger v.2.0.1 (https://www.10xgenomics.com/) and analyzed with the python package Scanpy v.1.7.1 (https://scanpy-tutorials.readthedocs.io/en/latest/index.html). Gene expression signatures were extracted using our SignatureAnalyzer algorithm available on GitHub (https://github.com/broadinstitute/getzlab-SignatureAnalyzer). Other analysis code will be uploaded to GitHub prior to publication.

## Conflict(s) of Interest

F.A. is an inventor on a patent application related to SignatureAnalyzer-GPU; G.G. receives research funds from IBM & Pharmacyclics, and is a founder, consultant, and has privately held equity in Scorpion Therapeutics; G.G is also an inventor on patent applications filed by the Broad Institute related to MSMuTect, MSMutSig, POLYSOLVER, SignatureAnalyzer-GPU, and MSIDetect; I.M.G. is a Consultant for AbbVie, Adaptive, Bristol Myers Squibb, Celgene Corporation, Cellectar, CohBar, Curio Science, Dava Oncology, Genentech, Huron Consulting, Karyopharm, Magenta Therapeutics, Menarini Silicon Biosystems, Oncopeptides, Pure Tech Health, Sognef, Takeda, and The Binding Site; an Advisor for Mind Wrap Medical, LLC; and an Advisor and Consultant for Amgen, Aptitude Health, GlaxoSmithKline, GNS Healthcare, Janssen, Pfizer, and Sanofi. I.M.G.’s spouse, William Savage MD, PhD, is CMO and equity holder of Disc Medicine (Private company, not publicly traded); N.J.H. is a consultant for Constellation Pharmaceuticals; D.S. is a consultant for ASAPP, has privately held equity in Curai and ASAPP, and receives research funds from Takeda and IBM; O.Z. is an employee at Constellation Pharmaceuticals.

## Author Contributions

Conception and design: R.B., N.J.H., R.S.P., D.S., I.M.G., G.G.

Collection and assembly of data: T.H.M., O.Z., H.E.K., D.F., N.J.H, S.K.A., R.S.P., R.B.

Data analysis and interpretation: R.B., N.J.H., J.B.A., F.A., M.C.S., R.S.P., D.S., I.M.G., G.G.

Funding acquisition: D.S., I.M.G., G.G.

Supervision: D.S., I.M.G., G.G.

Manuscript writing: R.B., N.J.H., J.B.A., M.M., F.A., D.S., I.M.G., G.G.

## Acknowledgements

Anna Justis, PhD, a medical writer employed by Dana-Farber Cancer Institute, supported the preparation of this manuscript. We would also like to thank Monica Agrawal, Michael Timonian and Mahshid Rahmat for their input on the manuscript. MMRF CoMMpass data used in analysis were generated as part of the Multiple Myeloma Research Foundation Personalized Medicine Initiatives (https://research.themmrf.org and www.themmrf.org). R.B. and D.S were supported by an ASPIRE award from The Mark Foundation for Cancer Research. G.G. is partially supported by the Paul C. Zamecnik Chair in Oncology at the Massachusetts General Hospital Cancer Center. This study was partially funded by the Multiple Myeloma Research Foundation (MMRF), Stand Up to Cancer (SU2C), and the Dr. Miriam and Sheldon G. Adelson Medical Research Foundation (AMRF).

---

**Extended Data Figure 1.**
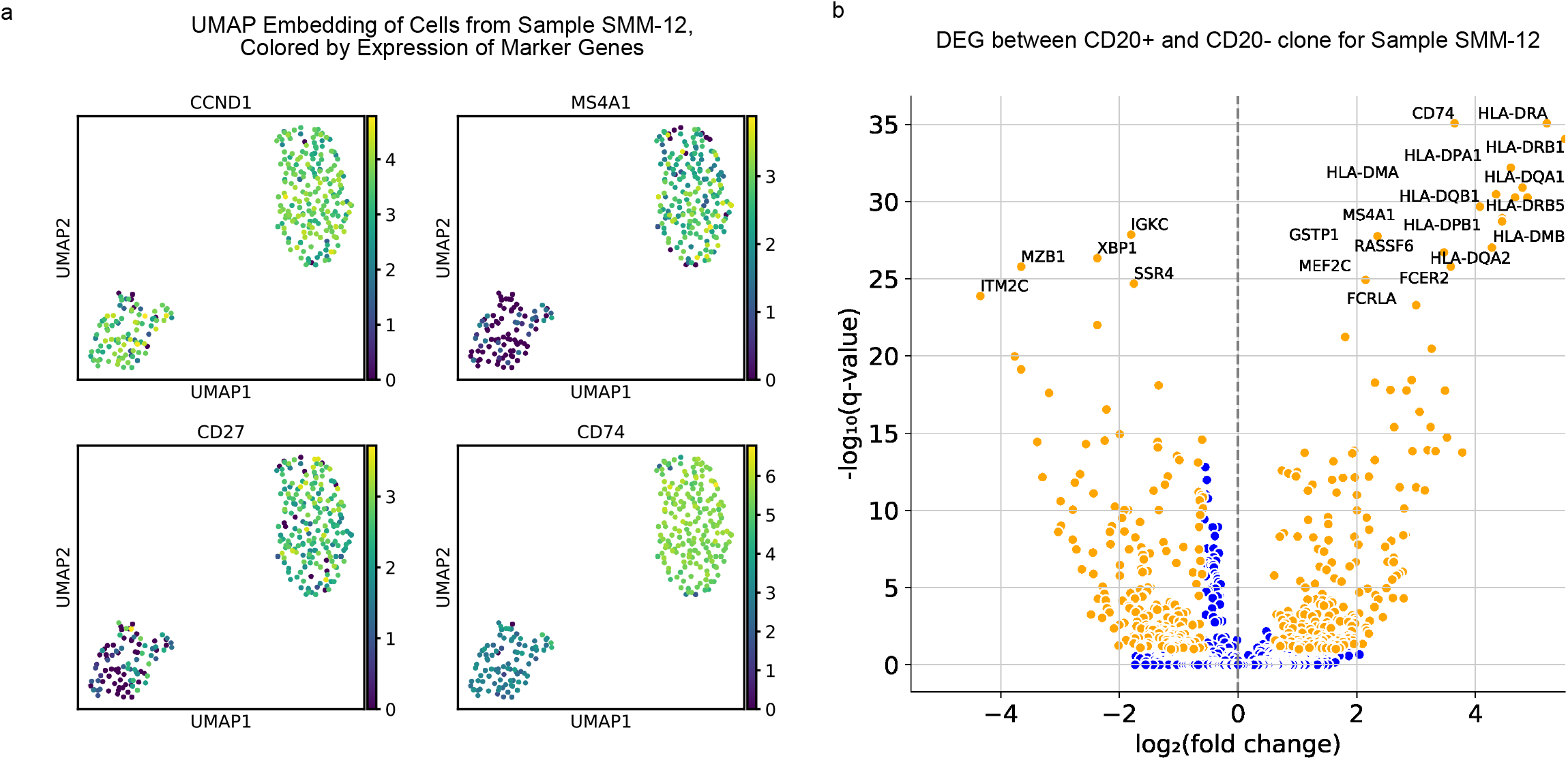
Sample SMM-12 has t(11;14) translocation and CD20+ subclone. **a,** UMAP embedding of cells from sample SMM-12, which has 190 cells belonging to a CD20+ subclone (cluster on right) out of 284 total cells. Both clusters express *CCND1*, signifying that both harbor a t(11;14) translocation, but the cluster on the right expresses higher levels of *CD20 (*also known as *MS4A1), CD27, CD74*. **b,** Volcano plot of DEGs between the CD20+ and CD20- subclones from sample SMM-12. Orange denotes a significant DEG (|log(fold change)| > log(1.5); q<0.1). The top genes, ranked by q-value, are annotated on the plot.

**Extended Data Figure 2.**
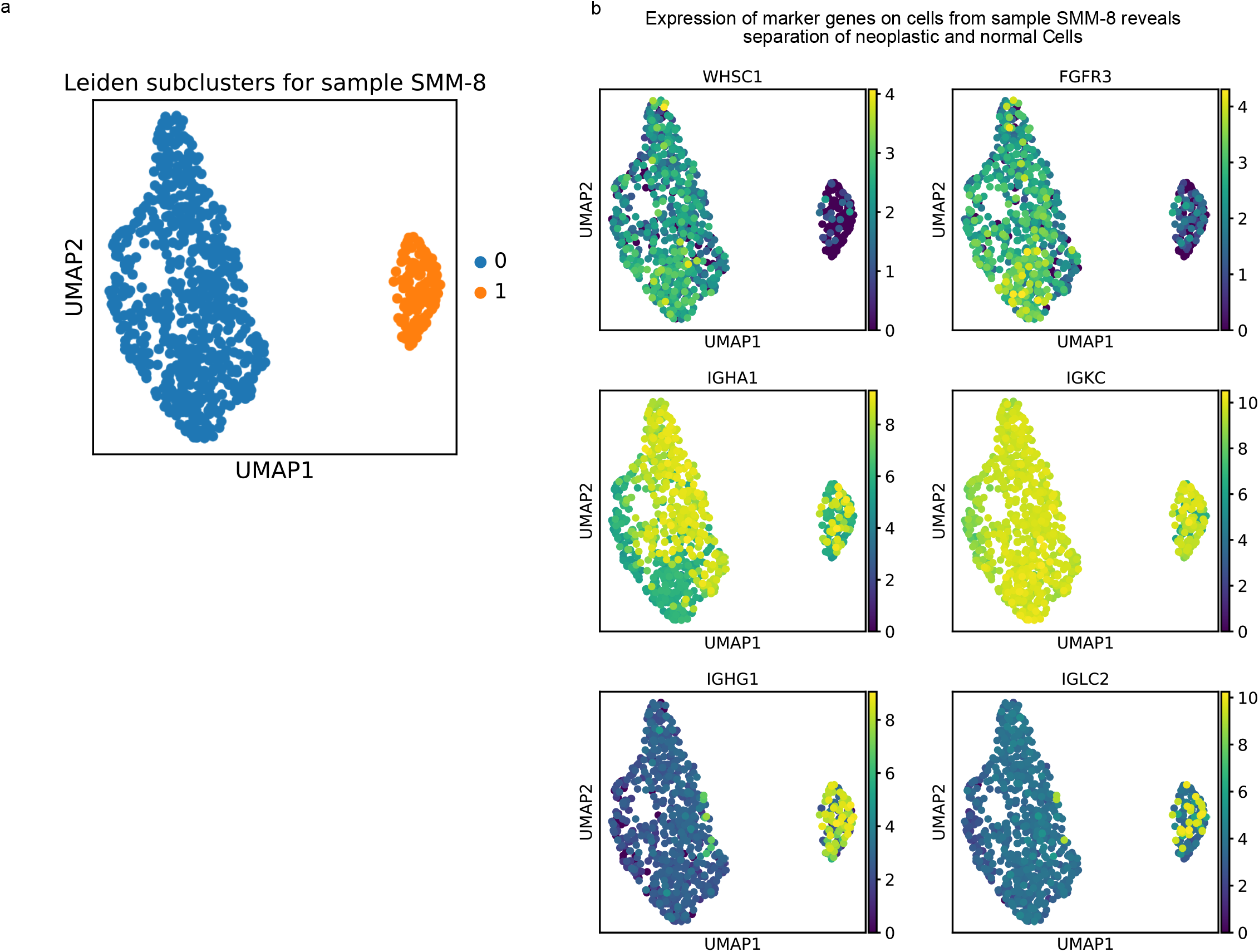
Example of subclustering approach for labeling normal and neoplastic plasma cells within each sample. **a,** UMAP plot showing subclustering result for representative sample SMM-8. Genes in immunoglobulin loci were not used in the computation of the UMAP embedding or Leiden clustering. **b,** Expression of genes used to determine that cluster 0 contains neoplastic cells and the cluster 1 contains normal cells: t(4;14) translocation associated genes *WHSC1* and *FGFR3* (first row); genes encoding the IgA-kappa immunoglobulin expressed on this patient’s monoclonal cells (second row); genes encoding non-clonal immunoglobulin components IgG (*IGHG1*) and the lambda light chain (*IGLC2*) (third row). We observe that cells in cluster 0 clonally express IgA-kappa genes, while cluster 1 contains a mixture of cells expressing IgA and IgG heavy chains and kappa and lambda light chains. We performed a similar analysis for every patient sample in our cohort.

**Extended Data Figure 3.**
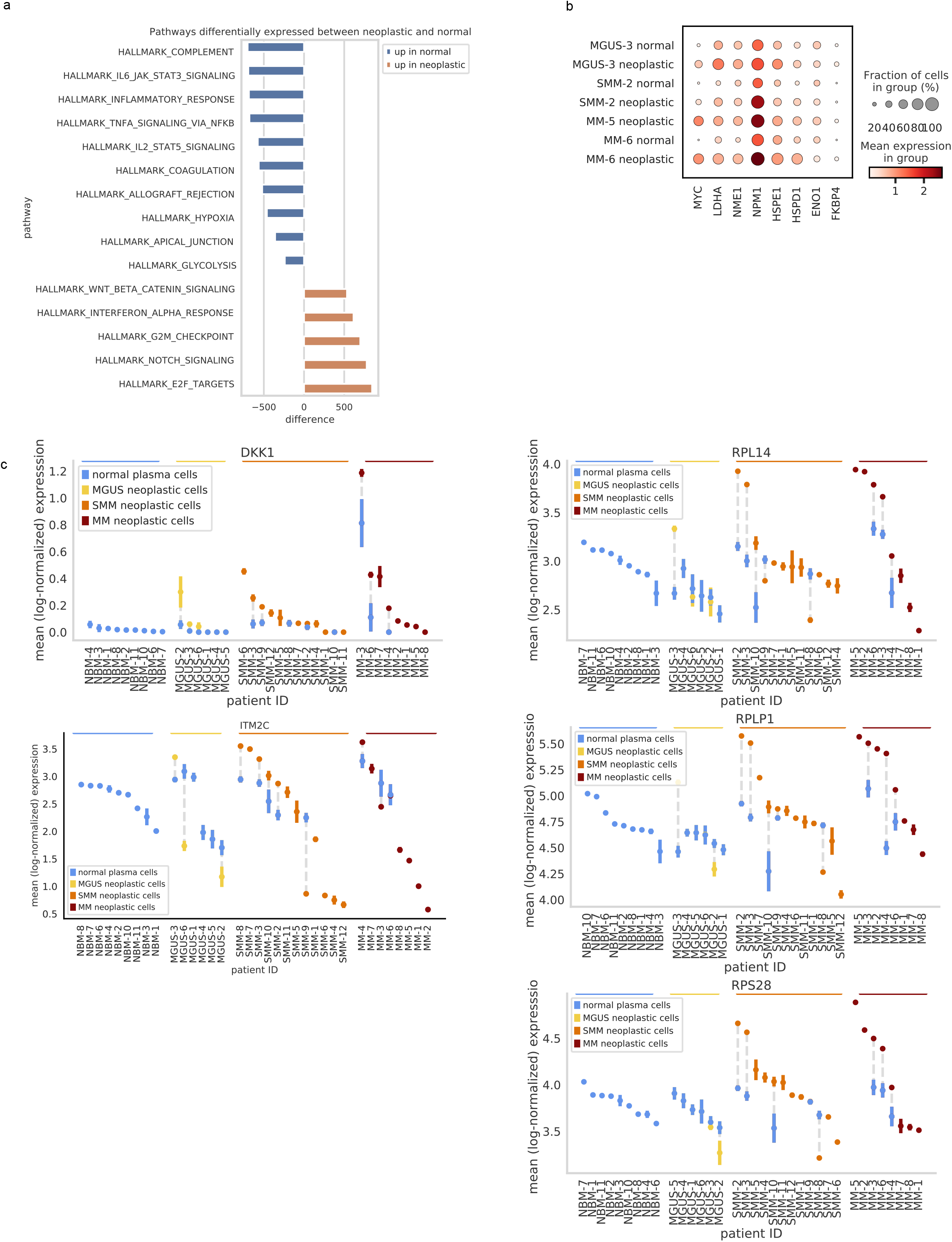
Additional visualizations of transcriptional differences detected between normal and neoplastic cells. **a,** MSigDB hallmark genesets differentially enriched in neoplastic samples compared to normal (t-test, q<0.1). The difference between the mean enrichment among neoplastic vs. normal samples is plotted on the x-axis. **b,** Expression of *MYC* and the MYC activation signature genes reported in Chng et al., 2011 in samples with significant DE of *MYC* by within-patient DE (MGUS-3, SMM-2, MM-6), as well as in MM-5, which was not included in the within-patient DE due to 100% purity, but had high levels of *MYC* comparable to MM-6. Cells are grouped by normal/neoplastic labeling as well as sample ID (y-axis). Color intensity corresponds to the mean expression of the gene in each group, and dot size corresponds to the fraction of cells in the group that express the gene. **c,** Mean expression ± s.e.m. of selected DEGs across the normal and neoplastic portions of samples at different disease stages.

**Extended Data Figure 4.**
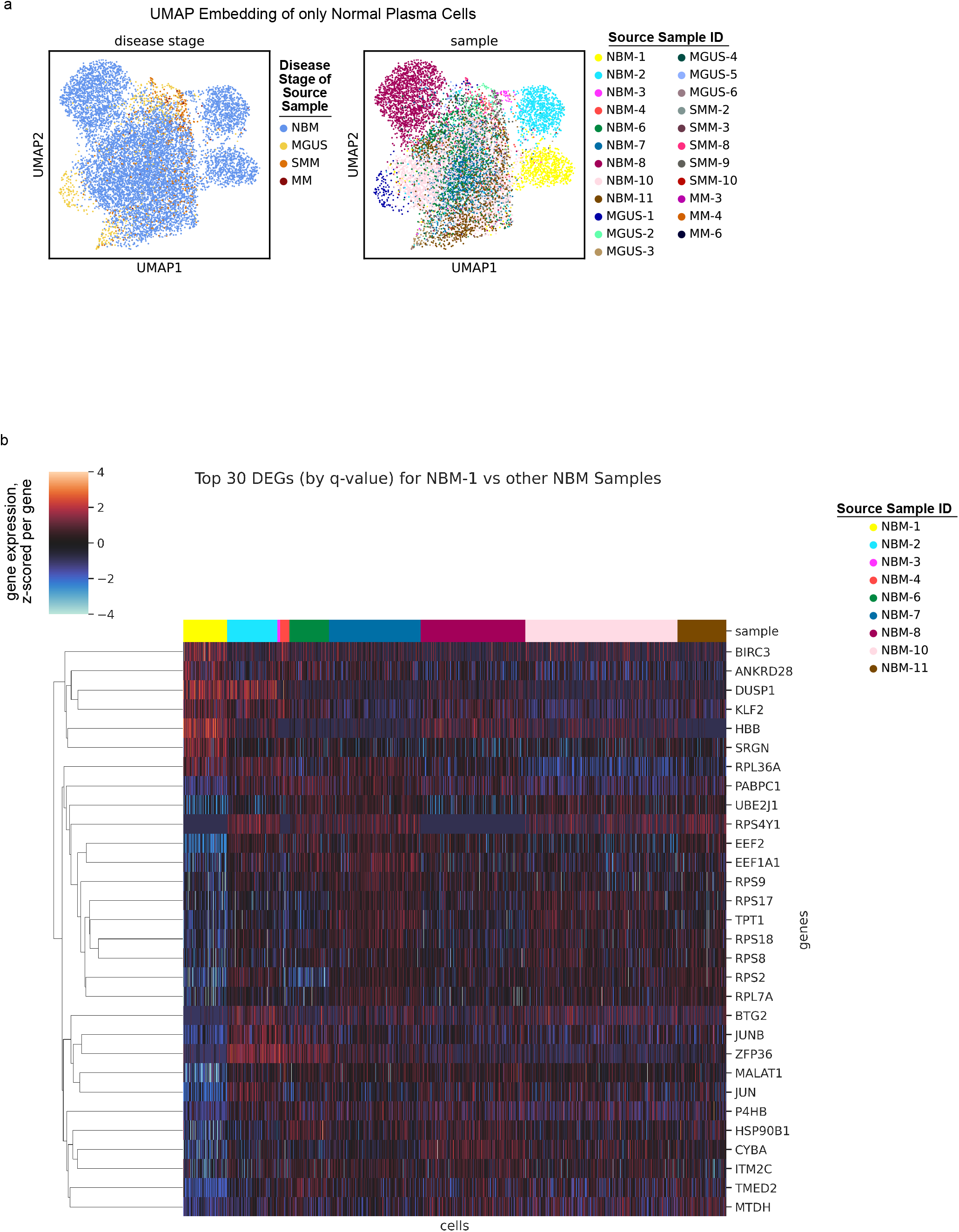
Inter-patient variation among healthy cells. **a,** Inter-patient variation exists even among normal plasma cells (even among those from NBM donors), as observed in these UMAP embeddings containing only cells labeled as normal. The normal plasma cells on these plots are colored by the disease stage (left) and sample ID (right) of the patient from which they were extracted. **b,** As further evidence of inter-patient variation among NBM samples, we found that comparing one NBM sample against the others returns many DEGs. As an example, comparing cells from patient NBM-1 to those from other NBM patients returned 1,061 DEGs (|log(fold change)| > log(1.5); q<0.1). Here, we visualize the top 30 DEGs (by q-value) for NBM-1 vs. other NBMs. Each column represents the gene expression of a single cell, and cells are ordered by the samples from which they originated. Expression is z-scored row-wise.

**Extended Data Figure 5.**
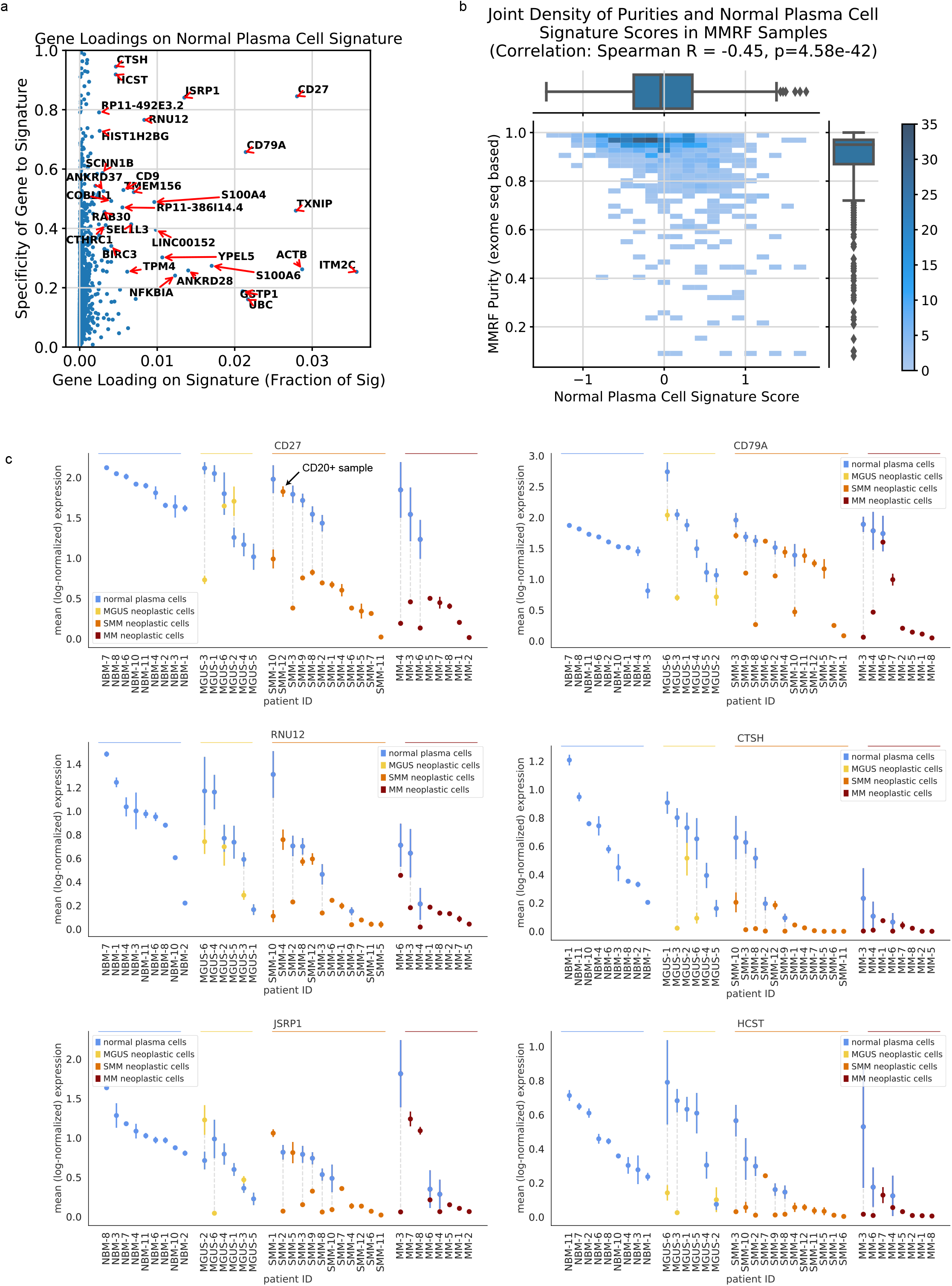
Expression of top genes in normal plasma cell signature and validation in bulk MMRF data. **a,** Contributions of individual genes to the ‘normal plasma cell signature.’ The x-axis represents a gene’s loading on the signature, i.e. its value in the W matrix, and the y-axis represents a gene’s specificity to the signature (see Methods). **b,** In bulk samples from the MMRF dataset, we observe a significant negative correlation between sample purity and ‘normal plasma cell signature’ score. The joint density of purity and signature score is plotted, with the intensity of color indicating the number of samples at a given part of the distribution (see color bar). Boxplots representing the marginal distributions of signature scores and purities are plotted along the top and right, respectively (center line, median; box limits, upper and lower quartiles; whiskers, 1.5x interquartile range; points, outliers). **c,** Mean expression ± s.e.m. of top genes from the ‘normal plasma cell signature’ in neoplastic and normal plasma cell portions of samples. We see that expression of the top genes from the signature are generally also downregulated in neoplastic cells as compared to normal plasma cells at all stages of disease. For *CD27*, the one notable exception is sample SMM-12, which has a CD20+ phenotype.

**Extended Data Figure 6.**
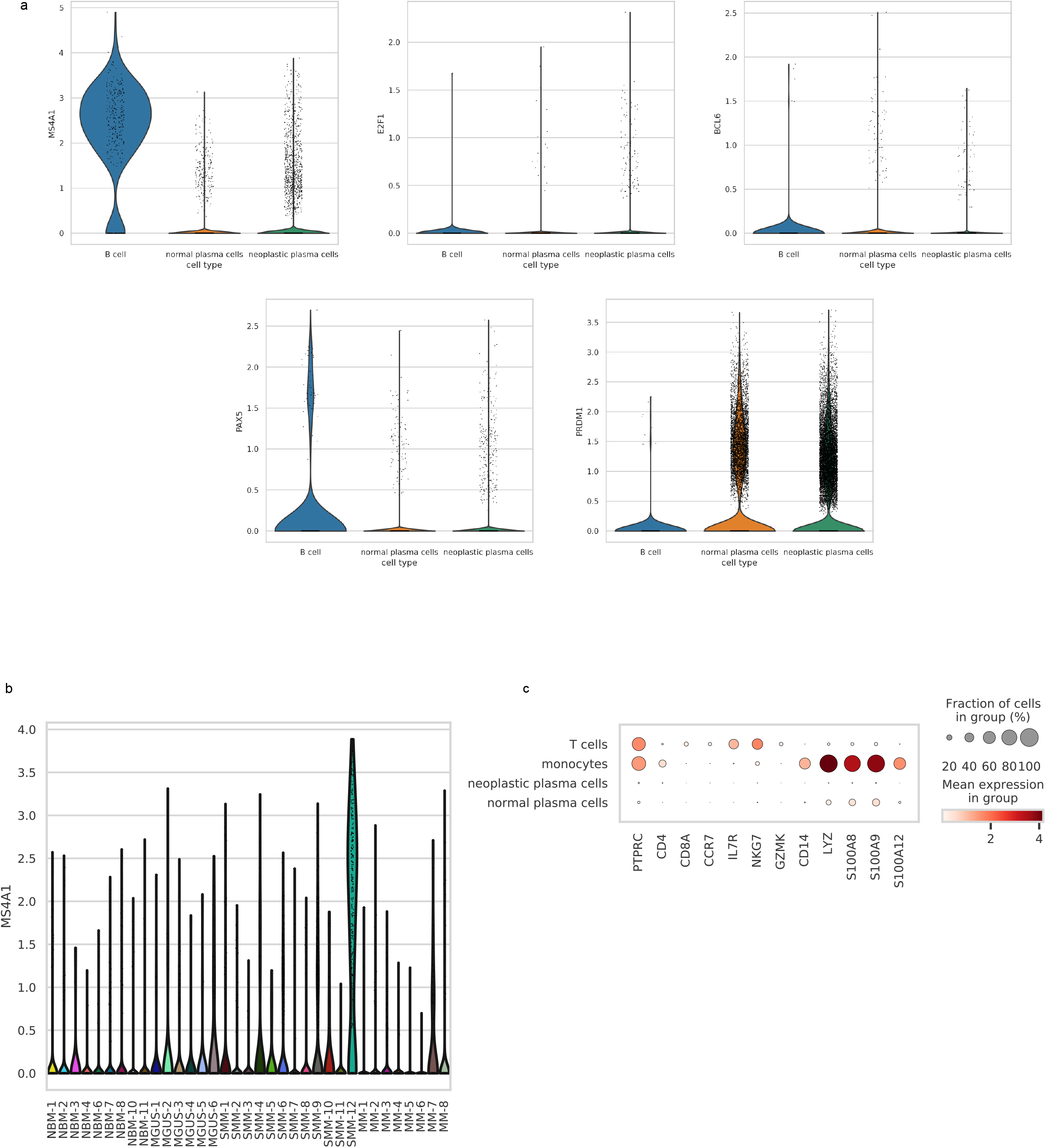
No contamination from B cells, T cells, or monocytes in CD138+ cells. **a,** B cell surface marker *CD20* (also known as *MS4A1)* and B cell transcription factors *E2F1, PAX5,* and *BCL6* are virtually not expressed in normal or neoplastic CD138+ cells (with the exception of *CD20* expression in SMM-12; see **(b)**), while they are expressed on the B cells which were removed from our data as part of QC. The plasma cell transcription factor *PRDM1* is expressed on CD138+ cells, but not B cells. Violin plots show distribution of expression over cells in each group. **b,** Expression of *CD20* on CD138+ cells is largely driven by patient SMM-12, who has a CD20+ MM phenotype. **c,** T cell and monocyte marker genes are hardly expressed in our CD138+ normal or neoplastic cells, while they are expressed in the T cell and monocyte populations which we removed from our data as part of QC. While there are low levels of monocyte marker genes among normal PCs, indicating possible low levels of ambient contamination in some normal samples, these genes are not expressed in neoplastic cells, indicating that neoplastic PCs (i.e. the PCs plotted in Figure 4a for comparison with monocytic populations) are uncontaminated. Color intensity corresponds to the mean expression of the gene in each group, and dot size corresponds to the fraction of cells in the group that express the gene.

**Extended Data Figure 7.**
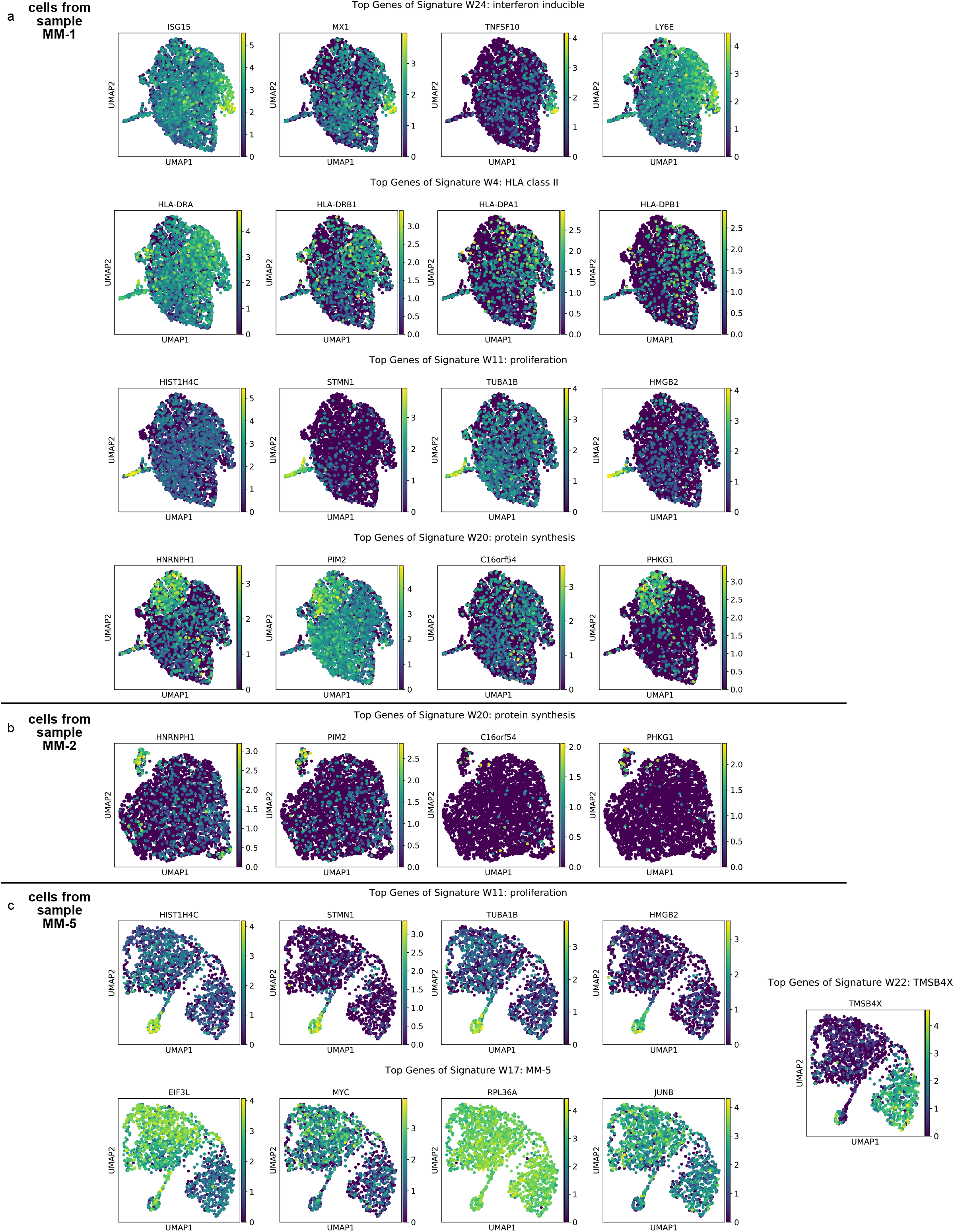
Genes are heterogeneously expressed within tumor samples. UMAP embeddings of cells from samples MM-1 **(a)**, MM-2 **(b)**, and MM-5 **(c)**, colored by expression of the top 4 genes from each signature that was highlighted as heterogeneously active within that patient in Fig. 4c. Since signature 22 was a single-gene signature, we only plotted the expression of the top gene (*TMSB4X)*.

## Notes

### Author Declarations

These studies were approved by the Dana-Farber Cancer Institute IRB. Informed consent was obtained from all patients and healthy volunteers in accordance with the Declaration of Helsinki protocol (fifth revision from 2000 with Clarifications of Articles 29, 30 (20022004), and the most recent iteration from 2013). MGUS and SMM patient samples were collected for a clinical trial, clinicaltrial.gov identifier NCT02269592.

